# Brain connectivity and motor improvements after ballet intervention in multiple sclerosis: pilot

**DOI:** 10.1101/2021.03.10.21252757

**Authors:** Paul B. Camacho, Brad P. Sutton, Citlali López-Ortiz

## Abstract

**Background and Purpose:** A pilot study to determine feasibility of detecting changes in structural connectivity (SC) and resting-state functional connectivity (RSFC) occur alongside motor improvements after participation in the Targeted Ballet Program (TBP) in adults with relapsing-remitting multiple sclerosis (RRMS).

**Methods:** Five participants (four female) with RRMS between the ages of 38-64 with the following characteristics at baseline: Expanded Disability Status Scale 2.0-6.0, International Cooperative Ataxia Rating Scale (ICARS) > 7, Symbol-Digit Modality Test > 22, and no relapses or initiation of medications indicated to affect mobility within the past 30 days. Participants were asked to complete 12 weeks (one hour, twice per week) of the TBP. Magnetic resonance imaging data was collected pre- and post-intervention for SC and RSFC network analysis.

**Results:** Increases in two RRMS-related graph theoretical measures (mean strength and mean clustering coefficient) for RSFC (p < 0.05) are detectable alongside significant reduction in ataxia (ICARS: p = 0.01012, Smoothness Index: p = 0.04995), and increase in balance (Mini-BESTest: p = 0.01474) following participation in the well-tolerated TBP.

**Discussion and Conclusions:** Significant increases in mean strength and mean clustering coefficient of RSFC suggest functional neurological improvements after participation in the TBP. The relationship between these network changes and clinical improvements in balance and amelioration of ataxia after participation in the TBP requires a larger randomized-controlled clinical trial of the TBP in persons with RRMS.

## Introduction

Multiple sclerosis (MS) is an autoimmune-mediated disease with brain demyelination and axonal loss causing impaired mobility and ataxia, which affect an estimated 75% and 80% of persons with MS respectively ^1-3^. Ataxia presents as varied combinations of unsteady gait, limb trajectory errors, trunk instability, motor sequencing and timing errors, speech impairment, decreased tone, abnormal stretch reflexes, and body or head tremor ^4^. An estimated 900,000 persons in the U.S. suffer from MS, which has no known cure ^5^ or effective pharmacological treatment for restoring mobility and decreasing ataxia ^6,7^. Pharmacological treatment of MS consists of disease-modifying drugs that slow down disease progression by targeting immune system function. However, these drugs do not induce remyelination of the CNS and, therefore, loss of motor and sensory functions persists.

Studies in mice and in healthy adult humans suggest that myelination is required for motor learning ^8^. Learning complex motor tasks – such as juggling, balancing on an unstable board, rhythmically cued finger movements, and dance – has been shown to increase white matter integrity and structural connectivity (SC) in fiber tracts in the cerebellum and areas involved in visuo-motor coordination and intra- and interhemispheric communication in both healthy adults and after stroke ^8-17^. Resting-state functional connectivity (RSFC) ^18,19^ analyses have shown changes in magnetic resonance imaging (MRI) in healthy adults due to bimanual visual tracking training and musical performance training. However, it is not yet known whether learning complex motor movements causes similarly significant changes in brain connectivity in persons with MS.

A classical ballet-based program for complex motor learning delivered in a group setting, targeted to improve balance and reduce ataxia in persons with MS has shown positive results beyond those of other physical rehabilitation interventions ^20^. Regarding dance research, evidence shows positive training effects of classical ballet on balance^21^, postural control^22^, joint position sense^23^, and a shift from vision to proprioception in the coordination of movements^24,25^ in healthy adults, beyond the existing evidence for the benefits of social dance forms (e.g. salsa and ballroom dancing) ^23-32^. The foundation of the targeted ballet program (TBP) lies in complex motor learning intrinsic to western classical ballet training ^20^. Motor learning in ballet training organizes movement and its instruction hierarchically while requiring active allocation of proprioceptive, auditory, visuospatial, emotional, and attentional resources. Classical ballet training hierarchy is based on a set of postures organized in anatomical Cartesian planes. Trained ballet postures are akin to letters in the alphabet. Learning complexity evolves by connecting postures to create movement words and phrases^20^. This approach to movement composition resembles theories of motor control based in the compositionality of dynamic primitives: movements that connect consecutive postures may be thought of as the dynamic ballet primitives ^33-35^. By modulating the dynamics of movement phrases, specific movement qualities such as smoothness and static and dynamic equilibrium are explicitly trained. This process is amenable to the training of movement patterns for transfer to everyday functional mobility, thereby unlocking ballet’s potential for movement rehabilitation. Movement narratives with meaning during training enhance motivation and cognitive engagement during complex motor learning.

In a pilot study, this TBP was well-tolerated and showed evidence that it leads to improvements in motor measures in persons with relapsing-remitting MS (RRMS) ^20^. Prior to this work, the largest improvements due to physical activity interventions in MS were 11.5% for balance and 15.2% for a timed 10-meter walk test ^36,37^. TBP increased Mini-Balance Evaluation Systems Test (Mini-BESTest) scores by 42% (p = 1e-4, Cohen’s *d* = 1.2) and decreased International Cooperative Ataxia Rating Scale (ICARS) scores by 58% (p = 7.11e-5, Cohen’s *d* = 2.6) ^20^. However, we did not collect data on concurrent brain connectivity changes.

Brain imaging of SC before and after participation in TBP may provide evidence of neuroplasticity underlying motor improvements in persons with MS. Along with SC, RSFC measures the effective active communication between regions ^38^. RSFC correlates with SC ^39^, but may provide differential sensitivity to indirectly connected networks in MS. Previous research supports the potential of this method in MS. RSFC has been shown to be reproducible in patients with stable RRMS according to a recent study of 20 RRMS patients compared with 14 healthy controls ^40^. Due to simultaneous use of multiple sensory modalities and cognitive networks, we expect increases in RSFC network measures after participating in the TBP.

## Methods

This research was approved by the Institutional Review Board at the University of Illinois at Urbana-Champaign in accordance with the Declaration of Helsinki. All participants gave voluntary informed consent after approval for participation. This preliminary study is registered in ClinicalTrials.gov under the trial identifier NCT04073940 (https://clinicaltrials.gov/ct2/show/NCT04073940).

### Participants and Setting

Inclusion criteria were: age 18-64 years, informed consent, confirmation of RRMS diagnosis and movement training approval from participant’s neurologist, Expanded Disability Status Scale scores of 2.0-6.5, relapse-free for 30 days, and absence of other central nervous system conditions. Exclusion criteria were: Symbol Digit Modalities Test < 23, pregnancy, education level < 8th grade, change in disease modifying therapy within 6 months, initiating medications that influence mobility within 30 days, orthopedic or musculoskeletal conditions, and contra-indications for MRI. Recruitment began upon IRB approval on June 6^th^ of 2018 and ended on September 19^th^ of 2019.

Pre-intervention assessments began on August 20^th^ of 2019 and ended on September 19^th^ of 2019. Post-intervention assessments were performed between December 3^rd^ of 2019 and December 13^th^ of 2019. All motor assessments and TBP classes were performed at the University of Illinois at Urbana-Champaign in the ballet studio of the Neuroscience of Dance in Health and Disability Laboratory. MRI data was collected at the Biomedical Imaging Center of the Beckman Institute for Advanced Science and Technology at the University of Illinois at Urbana-Champaign. The University of Illinois at Urbana-Champaign is located in a semi-rural community.

### Intervention

Participants were asked to perform 12 weeks (one hour, two days/week) of group TBP classes led by a Bolshoi-certified ballet instructor (CLO). Weekly make-up classes were made available to account for pre-existing time conflicts expressed by multiple participants. TBP classes were taught from August 28^th^, 2019 to December 2^nd^, 2019. Every class consisted of a seated warm up and ballet technique (20 min), followed by exercises using ballet *barres* (15 min), exercises across the floor (20 min), and a cool-down (5 min) ^20^. The seated warm-up focuses on articulating feet and hands, with progression from distal to proximal movements throughout the body, emphasizing selective control of each joint. As participants gain selective joint control, the warm-up transitions to smoothly connected, whole-body movements that link proximal to distal joints and vice versa. During the seated ballet section of the TDP, participants execute movements starting with the with the lower limbs in parallel position and progressing to first position of the feet with external rotation at the hips, from which they perform exercises in an adapted manner. The seated exercises were developed to provide the motor coordination foundations for the ballet *barre* and across the floor exercises. Movement adaptations focus on maintaining key characteristics of each ballet movement ^41^. Exercises were selected to target movement deficits observed in the group. Whenever possible, participants performed all movements to the front, side, and back directions. Movements were performed in the allowable range of motion of the joints of each participant, to minimize the possibility of injury. All instruction conformed to the principles of classical ballet technique and focus on control of the trunk with awareness of all joints in postures and movements. Movement difficulty was comparable to the Royal Academy of Dance and the Cecchetti Council of America Ballet I-II Syllabi ^42,43^. The instructor and assistants ensured that participants performed the intervention with the equivalent of the moderate level of the Borg Rating of Perceive Exertion Scale^44^, with focus on motor learning over aerobic or muscular exertion. University-level graduate and undergraduate trained assistants at a ratio of 1:1 to 2:1 facilitated fidelity in the execution of the movement presented and adherence to the prescribed movement. Movements were performed in time with piano recordings from Dmitri Roudnev’s Ballet Class Music Series^45^

## Main Outcome Measures

### Feasibility

Recruitment and retention rates were calculated as basic measures of trial feasibility. A retention rate of 80% or better was considered feasible due to similar rates observed in prior MS studies in our setting. Adherence to protocol was determined by the number of hours of TBP instruction time completed by each participant in comparison to the 24 hours of prescribed instruction time. Other dance forms have been well-tolerated in MS ^46^, thus we expected the TBP to be well-tolerated as seen in our prior pilot study ^20^. Tolerability was measured by the ability of participants to complete each TBP class without related adverse events or issues of thermoregulation preventing participation. Testing burden was deemed feasible if participants could complete all testing measures within the prescribed testing window without requesting adjustment to protocol. Detection of significant motor improvements and changes in SC and RSFC metrics served as critical measures of effect-detection feasibility.

### Motor function

We clinically assessed ataxia using ICARS, the leading comprehensive clinical measure of ataxia in MS with strong inter-evaluator reliability and validity ^47-49^. Higher ICARS scores indicate increased ataxia. We assessed balance ability using the Mini-BESTest, which has been validated in MS ^50-52^. Lower scores for the Mini-BESTest indicate better control of balance. Using a Qualisys motion capture system (Qualisys AB, Goteborg, Sweden), we assessed ataxia quantitatively using the spectral arc length metric yielding a bilateral smoothness index (s-index) ^20,53^,54 calculated using the SpectralArcLength.m function from https://github.com/siva82kb/smoothness ^53^.

### MRI Connectivity

We calculated three graph theory metrics (GTMs): mean strength, global efficiency, and mean clustering coefficient that have been used previously in MS to characterize disruptions in SC and RSFC in relation to sensorimotor function ^55-58^. We expected to detect increases in these GTMs for both SC and RSFC based on large increases in associated motor function seen in our prior pilot of the TBP ^20^.

### Acquisition

For diffusion-weighted imaging (DWI) on the Siemens 3 T Prisma (Siemens, Erlangen, Germany) using a 64-channel head coil, the CMRR multiband sequence (https://www.cmrr.umn.edu/multiband/; ^59,60^) was used for both the fMRI and DWI data achieving whole brain coverage with 2.5 mm isotropic resolution, multiband factor of 4. DWI data had 64 diffusion encoded directions in 2 shells (b-value = 1500 and 3000 s/mm^2^). Blood-oxygen-level dependent (BOLD) fMRI data (TR 2 s, TE 40 ms, 96 slices of 2.5 mm slice thickness, flip angle 52°) was collected for a 9-minute acquisition using a visual fixation point as performed for MS in Bollaert et al ^61^. Standard structural data including a 0.9 mm isotropic T1-weighted 3D MPRAGE (TR 2.3 s, TE 2.32 ms, TI 900 ms, flip angle 8°) and 1.0 mm isotropic T2-weighted flow attenuated inversion recovery (FLAIR) acquisition were acquired as in our prior RRMS research ^62^.

### Preprocessing for Resting-State Functional Connectivity

Structural and fMRI data were preprocessed with fMRIPrep 20.0.6 ^63^, based on Nipype ^64^ 1.4.2. T1-weighted (T1w) volumes were corrected for intensity non-uniformity using N4BiasFieldCorrection ^65^ and skull-stripped using antsBrainExtraction.sh v2.1.0 using the NKI template. Brain surfaces were reconstructed using FreeSurfer recon-all v6.0.1 ^66^. Spatial normalization to the ICBM 152 Nonlinear Asymmetrical template version 2009c ^67^ was performed through nonlinear registration with antsRegistration ^68^.

Functional data was slice time corrected using 3dTshift from AFNI v16.2.07 ^69^ and motion corrected using mcflirt ^70^. Motion correcting transformations, field distortion correcting warp, BOLD-to-T1w transformation and T1w-to-template (MNI) warp were concatenated and applied using antsApplyTransforms using Lanczos interpolation.

### Resting-State Functional Connectivity

RSFC processing was performed using the 36 parameter confound regression and motion de-spiking pipeline design (see github.com/PennBBL/xcpEngine/blob/master/designs/fc36p_despike.dsn) in xcpEngine ^71^.Pearson’s correlations were then quantified for regions of interest in the Automated Anatomical Labeling Atlas ^72^. Network-based statistics ^55,56^ were calculated for the whole-brain network comprised of these regions of interest using the Brain Connectivity Toolbox (BCT) ^55,73^.

### Preprocessing for Structural Connectivity

Preprocessing was performed using *QSIPrep* 0.12.2, which is based on *Nipype* 1.5.1 ^64^. Many internal operations of QSIPrep use nilearn ^74^ and DIPY ^75^. The T1-weighted (T1w) image was corrected for intensity non-uniformity (INU) using N4BiasFieldCorrection^65^, and used as T1w-reference throughout the workflow. The T1w-reference was then skull-stripped using antsBrainExtraction.sh (ANTs 2.3.1), using OASIS as target template. Spatial normalization to the ICBM 152 Nonlinear Asymmetrical template version 2009c ^67^ was performed through nonlinear registration with antsRegistration^68^ (ANTs 2.3.1), using brain-extracted versions of both T1w volume and template. Brain tissue segmentation of cerebrospinal fluid (CSF), white-matter (WM) and gray-matter (GM) was performed on the brain-extracted T1w using FAST^76^ (FSL 6.0.3:b862cdd5). The DWI time-series were resampled to ACPC, generating a *preprocessed DWI run in ACPC space*.

### Structural Connectivity

Reconstruction was performed using *QSIprep* 0.12.2, which is based on *Nipype* 1.5.1 ^64^ Gorgolewski et al. (2018). Multi-tissue fiber response functions were estimated using the Dhollander algorithm. Fiber orientation distributions (FODs) were estimated via constrained spherical deconvolution ^77,78^ (CSD) using an unsupervised multi-tissue method ^79^. Reconstruction was done using MRtrix3^80^. FODs were intensity-normalized using mtnormalize ^81^. Three GTMs describing network properties that reflect communication disruption caused by MS were calculated using the BCT ^55,73^.

### Statistical Analysis

Outcome differences were tested for normality using the Shapiro-Wilk Test (*p* < 0.05) and visual assessment of QQ-plots and boxplots ^82-84^. We then conducted single-tailed paired *t*-tests in R (R Core Team, Vienna, Austria) ^85^. Given that this was a single-center pilot study, feasibility measures were not statistically tested due to low generalizability of such statistics.

## Results

Twenty-one potential participants expressed interest in the study, six of whom were available to regularly attend the TBP classes and complete eligibility screening. Safety precautions related to COVID-19 prevented offering TBP classes for four potential participants. One potential participant was ineligible due to age outside of the range approved for inclusion. Five participants were eligible, provided informed consent, and completed the study: four female, one left-handed, two retired and three employed, three Caucasian, one Hispanic/Latinx, and one African-American. Participants had the following characteristics at baseline and hours of participation (see Table 1):

**Table 1.**
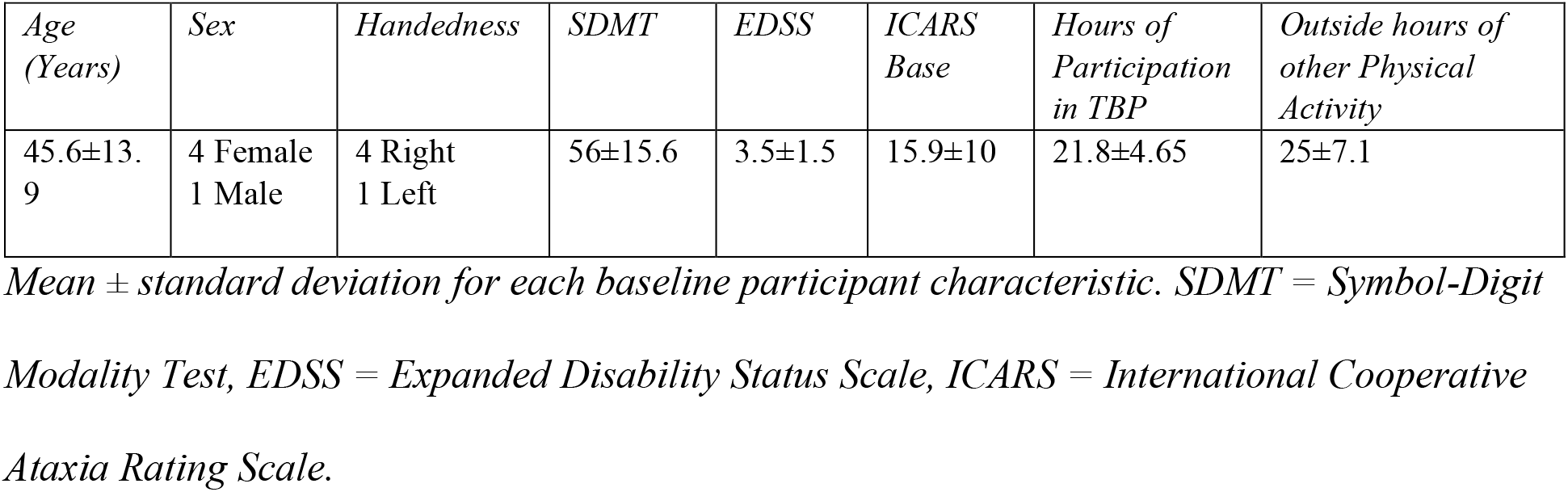
Baseline Participant Characteristics and Intervention Participation.

One participant stopped at 50% of the intervention duration due to an unrelated knee injury. However, this participant performed all post-intervention testing and is included in statistical analysis. This retention rate of 80% in the intervention is in line with previous studies on MS in our group and setting. The TBP was well-tolerated, with no adverse events related to participation and with successful thermoregulation of the dance studio and availability of trained assistants for comfortable completion of each class. All motor testing was completed within a two-hour block of testing and MRI scans were completed in less than one hour per session.

Paired t-tests showed reduction in ataxia (ICARS: *p* = 0.01012; bilateral s-index: *p* = 0.04995) and increased balance (Mini-BESTest: *p* = 0.01474). Mean strength and mean clustering coefficient increased for RSFC data (*p* < 0.05) (see Table 2). No significant changes were detected in structural connectivity analysis. Raw data for these outcomes are available upon request.

**Table 2.** Motor function outcomes.

| Participant | <u>**Motor Function**</u> |  |  |  |  |  |  |  |
| --- | --- | --- | --- | --- | --- | --- | --- | --- |
|  | *ICARS*<br>*Score*<br>*Pre* | *ICARS*<br>*Score*<br>*Post* | *ICARS*<br>*Posture*<br>*Gait*<br>*Pre* | *ICARS*<br>*Posture*<br>*Gait*<br>*Post* | *Mini-*<br>*BESTest*<br>*Score Pre* | *Mini-*<br>*BESTest*<br>*Score Post* | *Bilateral*<br>*s-index*<br>*Pre* | *Bilateral s-*<br>*index Post* |
| 1 | 10 | 8 | 2 | 1 | 23 | 26 | -79.7311 | -77.5589 |
| 2 | 33 | 31.5 | 12 | 7.5 | 16 | 18 | -101.2925 | -100.0835 |
| 3 | 9 | 4.5 | 5 | 1.5 | 22 | 26 | -85.6486 | -86.6932 |
| 4 | 17 | 16 | 2 | 2 | 23 | 23 | -88.6886 | -83.7198 |
| 5 | 10.5 | 6.5 | 3.5 | 0 | 25 | 27 | -89.4498 | -85.5882 |
| *Paired t-*<br>*test* | $p = 0.01012^*$ | | $p = 0.02128^*$ | | $p = 0.01474^*$ | | $p = 0.04995^*$ | |
| *Cohen's d*<br>*[95% CI]* | -1.67<br>[-4.55, 1.21] |  | -1.31<br>[-4.05, 1.42] |  | 1.48<br>[-1.32, 4.29] |  | 0.95<br>[-1.67, 3.57] |  |
| *Hedge's g*<br>*[95% CI]* | -1.21<br>[-3.18, 0.751] |  | -0.95<br>[-2.86, 0.951] |  | 1.08<br>[-0.854, 3.01] |  | 0.69<br>[-1.17, 2.55] |  |

For paired *t*-tests, \* denotes *p*-value < 0.05. ICARS = International Cooperative Ataxia Rating Scale, Mini-BESTest = Mini-Balance Evaluation Systems Test, *s*-index = smoothness index, 95% CI = 95% confidence interval. Participant numbers have been deidentified.

**Table 3.** Brain Connectivity Outcomes.

| Participant | Structural Connectivity |  |  | Resting-State Functional Connectivity |  |  |
| --- | --- | --- | --- | --- | --- | --- |
| | % $\Delta$ Global Efficiency | % $\Delta$ Mean Strength | % $\Delta$ Mean Clustering Coefficient | % $\Delta$ Global Efficiency | % $\Delta$ Mean Strength | % $\Delta$ Mean Clustering Coefficient |
| 1 | 0.05171 | 0.04623 | 0.05894 | -0.2194 | -0.1845 | -0.02534 |
| 2 | -0.05888 | -0.02900 | -0.03739 | 0.08331 | 0.2422 | 0.5232 |
| 3 | 0.005209 | 0.001800 | 0.01926 | 3.1845 | 2.907 | 0.8463 |
| 4 | 0.08198 | 0.07607 | 0.08625 | 1.236 | 1.042 | 0.2145 |
| 5 | -0.02414 | -0.02105 | -0.03118 | 1.639 | 1.5201 | 0.4386 |
| Paired t-test *p*-value | 0.639 | 0.750 | 0.749 | 0.0678 | 0.0491* | 0.0142* |
| Cohen's *d* [95% CI] | 0.172 [-2.32, 2.66] | 0.329 [-2.17, 2.83] | 0.332 [-2.17, 2.83] | 0.834 [-1.75, 3.42] | 0.961 [-1.66, 3.58] | 1.49 [-1.31, 4.31] |
| Hedge's *g* [95% CI] | 0.125 [-1.68, 1.93] | 0.239 [-1.57, 2.05] | 0.241 [-1.57, 2.05] | 0.607 [-1.24, 2.45] | 1.37 [-1.16, 2.56] | 1.09 [-0.85, 3.03] |

*For percent change t-tests and paired t-tests, \*\* denotes $p$ -value < 0.01, 95% CI = 95%*
*confidence interval. Participant numbers have been deidentified.*

## Discussion

Originally, additional rounds of the TBP were planned to begin in the Spring semester with rolling-enrollment allowing testing and the intervention to minimize conflict with Winter holidays and maximize participant recruitment and adherence. However, recruitment for this pilot study was limited by the ongoing COVID-19 pandemic. The TBP was well-tolerated and participants were able to complete all testing. The intervention retention rate of 80% is in line with prior MS research in our setting and was due to outside circumstances unrelated to the intervention. As was originally planned for this pilot, future trials will be scheduled to avoid holiday conflicts and provide a longer period of TBP class availability to maximize adherence and consistent intervention delivery.

Motor improvements seen here are consistent with our pilot study of TBP in females with RRMS ^20^, including for the first time a male with RRMS. While still a very large effect, the ICARS *Cohen’s d* = 1.67 was lower than the corresponding *Cohen’s d* = 2.6 from our pilot study of TBP ^20^. These smaller effect sizes were expected due to lower baseline levels of ataxia and ∼25% fewer intervention hours spent than in the prior study ^20^. This effect size is also limited by less than full completion of the intervention by one participant. Improvements in the bilateral s-index (*Cohen’s d* = 0.95) and Mini-BESTest (*Cohen’s d* = 1.48) were similar to the respective large and very large effect sizes seen previously for TBP ^20^. Sample size bias-corrected *Hedge’s g* ^86^ yielded large effect sizes for all clinical outcomes and a medium effect size for the s-index (see Table 2).

As expected, alongside motor improvements, we observed increases in mean strength and mean clustering coefficient in the RSFC data. These metrics have been shown to be lower in persons with MS than healthy controls ^57,58^. Thus, their increases suggest improvement in persons with MS, bringing these measures closer to those seen in healthy sensorimotor function ^57,58^. The extent to which each GTM represents disease response to intervention is limited by largely cross-sectional current literature.

The lack of expected significant increases in SC measures may be due to the small sample size of this preliminary study. Based on these results and accounting for the 80% intervention completion rate, a future longitudinal randomized-controlled trial with a repeated measures ANOVA statistical design would require recruitment of n = 212 participants achieve a significant change at power = 0.80 and α = 0.005 (Bonferroni-corrected for 10 simultaneous comparisons) in the GTMs of interest for SC that showed non-negligible changes in this pilot. The same statistical design would require recruitment of n = 20 participants for a trial to detect significant changes in GTMs for RSFC. However, we acknowledge that this estimation is limited in generalizability to longitudinal and multi-site trials due to the single-center, pre- and post-intervention design of this pilot.

With only five participants, statistical power of a small sample limits conclusions about the general RRMS population. We acknowledge that significance of outcome changes is limited by the need for appropriate correction for multiple simultaneous comparisons. However, the detection of within-participant differences in motor measures, mean strength, and mean clustering coefficient in RSFC supports a larger longitudinal study of the TBP in persons with RRMS. This would be a longitudinal randomized-controlled trial of the TBP in persons with RRMS, comparing the effects of the TBP on the aforementioned connectivity measures in SC and RSFC and motor outcomes to those of an intensity-matched version of the National Multiple Sclerosis Society Stretching for People with MS group class augmented by walking and lower extremity functional exercises known to improve mobility in persons with RRMS ^36,87^.

## Conclusions

The detection of significant increases in mean strength and mean clustering coefficient of resting-state functional connectivity coinciding with clinical improvements in balance and amelioration of ataxia after participation in the TBP supports a larger randomized-controlled clinical trial of the TBP in persons with RRMS. Effect sizes seen in this preliminary study may be skewed due to a small number of participants but remain large/medium after correction for small sample size. Although generalizability to multi-site randomized-controlled trials is limited, enrollment, retention rates, and well-tolerated intervention and testing suggest that a larger clinical trial is feasible.

## Supporting information

GPower Protocol Resting State Functional Connectivity

GPower Protocol StructuralConnectivity

R Statistical Analysis Report

## Data Availability

Data related to this study is available upon request to the authors

## Acknowledgements

We thank the Arnold and Mabel Beckman Foundation for financial support through the Beckman Institute for Advanced Science and Technology. We greatly appreciate the Biomedical Imaging Center and the Neuroscience of Dance in Health and Disability Laboratory staff, especially Andrea Rivera-Maza and Nina Crouchelli.

## Authors’ Roles

CLO conceived, organized, and executed the project with assistance from PBC and BPS. BPS designed the MRI acquisition and analysis strategy with assistance from PBC and CLO. PBC processed and analyzed data with design, review, and critique from CLO and BPS. PBC wrote the manuscript with critique and assistance from CLO and BPS.

## References

1. Hobart JC, Riazi A, Lamping DL, Fitzpatrick R, Thompson AJ. Measuring the impact of MS on walking ability: the 12-Item MS Walking Scale (MSWS-12). Neurology. Jan 14 2003;60(1):31–36.

2. Larocca NG. Impact of walking impairment in multiple sclerosis: perspectives of patients and care partners. The patient. 2011;4(3):189–201.

3. Mills RJ, Yap L, Young CA. Treatment for ataxia in multiple sclerosis. The Cochrane database of systematic reviews. Jan 24 2007(1):CD005029.

4. Brunberg JA, Expert Panel on Neurologic I. Ataxia. AJNR Am J Neuroradiol. Aug 2008;29(7):1420–1422.

5. Wallin MT, Culpepper WJ, Campbell JD, et al. The prevalence of MS in the United States: A population-based estimate using health claims data. Neurology. Mar 5 2019;92(10):e1029–e1040.

6. Vargas DL, Tyor WR. Update on disease-modifying therapies for multiple sclerosis. Journal of investigative medicine : the official publication of the American Federation for Clinical Research. Jun 2017;65(5):883–891.

7. Akaishi T, Nakashima I. Efficiency of antibody therapy in demyelinating diseases. International immunology. Jul 1 2017;29(7):327–335.

8. Lakhani B, Borich MR, Jackson JN, et al. Motor Skill Acquisition Promotes Human Brain Myelin Plasticity. Neural plasticity. 2016;2016:7526135.

9. Wittenberg GF, Richards LG, Jones-Lush LM, et al. Predictors and brain connectivity changes associated with arm motor function improvement from intensive practice in chronic stroke. F1000Research. 2016;5:2119.

10. Scholz J, Klein MC, Behrens TE, Johansen-Berg H. Training induces changes in white-matter architecture. Nature neuroscience. Nov 2009;12(11):1370–1371.

11. Taubert M, Draganski B, Anwander A, et al. Dynamic properties of human brain structure: learning-related changes in cortical areas and associated fiber connections. J Neurosci. Sep 1 2010;30(35):11670–11677.

12. Moore E, Schaefer RS, Bastin ME, Roberts N, Overy K. Diffusion tensor MRI tractography reveals increased fractional anisotropy (FA) in arcuate fasciculus following music-cued motor training. Brain Cogn. Aug 2017;116:40–46.

13. Burzynska AZ, Jiao YQ, Knecht AM, et al. White Matter Integrity Declined Over 6-Months, but Dance Intervention Improved Integrity of the Fornix of Older Adults. Front Aging Neurosci. Mar 16 2017;9.

14. Seehaus A, Roebroeck A, Bastiani M, et al. Histological validation of high-resolution DTI in human post mortem tissue. Frontiers in neuroanatomy. 2015;9:98.

15. Chang EH, Argyelan M, Aggarwal M, et al. The role of myelination in measures of white matter integrity: Combination of diffusion tensor imaging and two-photon microscopy of CLARITY intact brains. NeuroImage. Feb 15 2017;147:253–261.

16. Laule C, Leung E, Lis DK, et al. Myelin water imaging in multiple sclerosis: quantitative correlations with histopathology. Multiple sclerosis. Dec 2006;12(6):747–753.

17. Baer LH, Park MT, Bailey JA, Chakravarty MM, Li KZ, Penhune VB. Regional cerebellar volumes are related to early musical training and finger tapping performance. NeuroImage. Apr 1 2015;109:130–139.

18. Solesio-Jofre E, Beets IAM, Woolley DG, et al. Age-Dependent Modulations of Resting State Connectivity Following Motor Practice. Front Aging Neurosci. 2018;10:25.

19. Li Q, Wang X, Wang S, et al. Musical training induces functional and structural auditory-motor network plasticity in young adults. Human brain mapping. May 2018;39(5):2098–2110.

20. Scheidler AM, Kinnett-Hopkins D, Learmonth YC, Motl R, Lopez-Ortiz C. Targeted ballet program mitigates ataxia and improves balance in females with mild-to-moderate multiple sclerosis. PLoS One. 2018;13(10):e0205382.

21. Gerbino PG, Griffin ED, Zurakowski D. Comparison of standing balance between female collegiate dancers and soccer players. Gait Posture. Oct 2007;26(4):501–507.

22. Rein S, Fabian T, Zwipp H, Rammelt S, Weindel S. Postural control and functional ankle stability in professional and amateur dancers. Clin Neurophysiol. Aug 2011;122(8):1602–1610.

23. Ramsay JR, Riddoch MJ. Position-matching in the upper limb: professional ballet dancers perform with outstanding accuracy. Clinical rehabilitation. Jun 2001;15(3):324–330.

24. Thullier F, Moufti H. Multi-joint coordination in ballet dancers. Neuroscience letters. Oct 07 2004;369(1):80–84.

25. Golomer E, Dupui P. Spectral analysis of adult dancers’ sways: Sex and interaction vision Proprioception. Int J Neurosci. 2000;105(1-4):15–26.

26. Kattenstroth JC, Kalisch T, Holt S, Tegenthoff M, Dinse HR. Six months of dance intervention enhances postural, sensorimotor, and cognitive performance in elderly without affecting cardio-respiratory functions. Front Aging Neurosci. 2013;5:5.

27. Kiefer AW, Riley MA, Shockley K, et al. Lower-limb proprioceptive awareness in professional ballet dancers. Journal of dance medicine & science : official publication of the International Association for Dance Medicine & Science. Sep 2013;17(3):126–132.

28. Schmit JM, Regis DI, Riley MA. Dynamic patterns of postural sway in ballet dancers and track athletes. Experimental brain research. Jun 2005;163(3):370–378.

29. Lepelley MC, Thullier F, Koral J, Lestienne FG. Muscle coordination in complex movements during Jete in skilled ballet dancers. Experimental brain research. Nov 2006;175(2):321–331.

30. Jola C, Davis A, Haggard P. Proprioceptive integration and body representation: insights into dancers’ expertise. Experimental brain research. Sep 2011;213(2-3):257–265.

31. Hanggi J, Koeneke S, Bezzola L, Jancke L. Structural neuroplasticity in the sensorimotor network of professional female ballet dancers. Human brain mapping. Aug 2010;31(8):1196–1206.

32. Teixeira-Machado L, Arida RM, de Jesus Mari J. Dance for neuroplasticity: A descriptive systematic review. Neurosci Biobehav Rev. Jan 2019;96:232–240.

33. Arbib MA. Perceptual structures and distributed motor control. In: Brooks VB, ed. Handbook of Physiology -The Nervous System II. Motor Control. Bethesda, MD: American Physiological Society; 1981:1449-1480.

34. Sternad D, Marino H, Charles SK, Duarte M, Dipietro L, Hogan N. Transitions between discrete and rhythmic primitives in a unimanual task. Front Comput Neurosci. 2013;7:90.

35. Flash T, Hogan N. The coordination of arm movements: an experimentally confirmed mathematical model. J Neurosci. Jul 1985;5(7):1688–1703.

36. Tarakci E, Yeldan I, Huseyinsinoglu BE, Zenginler Y, Eraksoy M. Group exercise training for balance, functional status, spasticity, fatigue and quality of life in multiple sclerosis: a randomized controlled trial. Clinical rehabilitation. Sep 2013;27(9):813–822.

37. Scult MA, Fresco DM, Gunning FM, et al. Changes in Functional Connectivity Following Treatment With Emotion Regulation Therapy. Frontiers in behavioral neuroscience. 2019;13:10.

38. Biswal BB, Mennes M, Zuo XN, et al. Toward discovery science of human brain function. Proceedings of the National Academy of Sciences of the United States of America. Mar 09 2010;107(10):4734–4739.

39. Honey CJ, Sporns O, Cammoun L, et al. Predicting human resting-state functional connectivity from structural connectivity. Proceedings of the National Academy of Sciences of the United States of America. Feb 10 2009;106(6):2035–2040.

40. Pinter D, Beckmann C, Koini M, et al. Reproducibility of Resting State Connectivity in Patients with Stable Multiple Sclerosis. PLoS One. 2016;11(3):e0152158.

41. Vaganova A. Basic Principles of Classical Ballet. Russian Ballet Technique.. Second Edition ed: Dover; 1965.

42. Royal Academy of Dancing. Children’s Examinations Syllabus for Girls and Boys. Revised ed. London, UK: Royal Academy of Dancing; 1984.

43. The Cecchetti Council of America. Graded Lessons In Classical Ballet Technique Grade One. Revised 2009 ed: The Cecchitti Council of America; 1991.

44. Borg G. Ratings of perceived exertion and heart rates during short-term cycle exercise and their use in a new cycling strength test. International journal of sports medicine. Aug 1982;3(3):153-158.

45. Roudnev D.

46. Mandelbaum R, Triche EW, Fasoli SE, Lo AC. A Pilot Study: examining the effects and tolerability of structured dance intervention for individuals with multiple sclerosis. Disability and rehabilitation. 2016;38(3):218–222.

47. Storey E, Tuck K, Hester R, Hughes A, Churchyard A. Inter-rater reliability of the International Cooperative Ataxia Rating Scale (ICARS). Movement disorders : official journal of the Movement Disorder Society. Feb 2004;19(2):190–192.

48. Trouillas P, Takayanagi T, Hallett M, et al. International Cooperative Ataxia Rating Scale for pharmacological assessment of the cerebellar syndrome. The Ataxia Neuropharmacology Committee of the World Federation of Neurology. Journal of the neurological sciences. Feb 12 1997;145(2):205–211.

49. Salci Y, Fil A, Keklicek H, et al. Validity and reliability of the International Cooperative Ataxia Rating Scale (ICARS) and the Scale for the Assessment and Rating of Ataxia (SARA) in multiple sclerosis patients with ataxia. Mult Scler Relat Disord. Nov 2017;18:135–140.

50. Godi M, Franchignoni F, Caligari M, Giordano A, Turcato AM, Nardone A. Comparison of reliability, validity, and responsiveness of the mini-BESTest and Berg Balance Scale in patients with balance disorders. Physical therapy. Feb 2013;93(2):158–167.

51. King L, Horak F. On the mini-BESTest: scoring and the reporting of total scores. Physical therapy. Apr 2013;93(4):571–575.

52. Franchignoni F, Horak F, Godi M, Nardone A, Giordano A. Using psychometric techniques to improve the Balance Evaluation Systems Test: the mini-BESTest. Journal of rehabilitation medicine. Apr 2010;42(4):323–331.

53. Balasubramanian S, Melendez-Calderon A, Burdet E. A robust and sensitive metric for quantifying movement smoothness. IEEE transactions on bio-medical engineering. Aug 2012;59(8):2126–2136.

54. Balasubramanian S, Melendez-Calderon A, Roby-Brami A, Burdet E. On the analysis of movement smoothness. Journal of neuroengineering and rehabilitation. Dec 9 2015;12:112.

55. Rubinov M, Sporns O. Complex network measures of brain connectivity: uses and interpretations. NeuroImage. Sep 2010;52(3):1059–1069.

56. Bullmore E, Sporns O. Complex brain networks: graph theoretical analysis of structural and functional systems. Nature reviews. Neuroscience. Mar 2009;10(3):186–198.

57. Liu Y, Wang H, Duan Y, et al. Functional Brain Network Alterations in Clinically Isolated Syndrome and Multiple Sclerosis: A Graph-based Connectome Study. Radiology. Feb 2017;282(2):534–541.

58. Shu N, Duan Y, Xia M, et al. Disrupted topological organization of structural and functional brain connectomes in clinically isolated syndrome and multiple sclerosis. Scientific reports. Jul 12 2016;6:29383.

59. Auerbach EJ, Xu J, Yacoub E, Moeller S, Ugurbil K. Multiband accelerated spin-echo echo planar imaging with reduced peak RF power using time-shifted RF pulses. Magn Reson Med. May 2013;69(5):1261–1267.

60. Setsompop K, Cohen-Adad J, Gagoski BA, et al. Improving diffusion MRI using simultaneous multi-slice echo planar imaging. NeuroImage. Oct 15 2012;63(1):569–580.

61. Bollaert RE, Poe K, Hubbard EA, et al. Associations of functional connectivity and walking performance in multiple sclerosis. Neuropsychologia. Aug 2018;117:8–12.

62. Wetter NC, Hubbard EA, Motl RW, Sutton BP. Fully automated open-source lesion mapping of T2-FLAIR images with FSL correlates with clinical disability in MS. Brain and behavior. Mar 2016;6(3):e00440.

63. Esteban O, Markiewicz CJ, Blair RW, et al. fMRIPrep: a robust preprocessing pipeline for functional MRI. Nature methods. Jan 2019;16(1):111–116.

64. Gorgolewski K, Burns CD, Madison C, et al. Nipype: a flexible, lightweight and extensible neuroimaging data processing framework in python. Frontiers in neuroinformatics. 2011;5:13.

65. Tustison NJ, Avants BB, Cook PA, et al. N4ITK: improved N3 bias correction. IEEE transactions on medical imaging. Jun 2010;29(6):1310–1320.

66. Dale AM, Fischl B, Sereno MI. Cortical surface-based analysis. I. Segmentation and surface reconstruction. NeuroImage. Feb 1999;9(2):179–194.

67. Fonov V, Evans, AC; McKinstry, RC; Almli, CR; Collins, DL. Unbiased nonlinear average age-appropriate brain templates from birth to adulthood. NeuroImage. Jul 1 2009 2009;47(S102).

68. Avants BB, Epstein CL, Grossman M, Gee JC. Symmetric diffeomorphic image registration with cross-correlation: evaluating automated labeling of elderly and neurodegenerative brain. Medical image analysis. Feb 2008;12(1):26–41.

69. Cox RW. AFNI: software for analysis and visualization of functional magnetic resonance neuroimages. Computers and biomedical research, an international journal. Jun 1996;29(3):162–173.

70. Jenkinson M, Bannister P, Brady M, Smith S. Improved optimization for the robust and accurate linear registration and motion correction of brain images. NeuroImage. Oct 2002;17(2):825–841.

71. Ciric R, Wolf DH, Power JD, et al. Benchmarking of participant-level confound regression strategies for the control of motion artifact in studies of functional connectivity. NeuroImage. Jul 1 2017;154:174–187.

72. Tzourio-Mazoyer N, Landeau B, Papathanassiou D, et al. Automated anatomical labeling of activations in SPM using a macroscopic anatomical parcellation of the MNI MRI single-subject brain. NeuroImage. Jan 2002;15(1):273–289.

73. Fleischer V, Radetz A, Ciolac D, et al. Graph Theoretical Framework of Brain Networks in Multiple Sclerosis: A Review of Concepts. Neuroscience. Apr 1 2019;403:35–53.

74. Abraham A, Pedregosa F, Eickenberg M, et al. Machine learning for neuroimaging with scikit-learn. Frontiers in neuroinformatics. 2014;8:14.

75. Garyfallidis E, Brett M, Amirbekian B, et al. Dipy, a library for the analysis of diffusion MRI data. Frontiers in neuroinformatics. 2014;8:8.

76. Zhang Y, Brady M, Smith S. Segmentation of brain MR images through a hidden Markov random field model and the expectation-maximization algorithm. IEEE transactions on medical imaging. Jan 2001;20(1):45–57.

77. Tournier JD, Yeh CH, Calamante F, Cho KH, Connelly A, Lin CP. Resolving crossing fibres using constrained spherical deconvolution: validation using diffusion-weighted imaging phantom data. NeuroImage. Aug 15 2008;42(2):617–625.

78. Tournier JD, Calamante F, Gadian DG, Connelly A. Direct estimation of the fiber orientation density function from diffusion-weighted MRI data using spherical deconvolution. NeuroImage. Nov 2004;23(3):1176–1185.

79. Dhollander TM, R; Raffelt, D; Connelly, A. Improved White Matter Response Function Estimation for 3-Tissue Constrained Spherical Deconvolution. Paper presented at: International Society of Magnetic Resonance in Medicine2019.

80. Tournier JS, R; Raffelt, D; Tabbara, R; Dhollander, T; Pietsch, M; Christiaens, D; Jeurissen, B; Yeh, CH; Connelly, A. MRtrix3: A Fast, Flexible and Open Software Framework for Medical Image Processing and Visualisation. NeuroImage. 2019;202:116–137.

81. Raffelt DD, T; Tournier, JD;Tabbara, R; Smith, RE; Pierre, E; Connelly, A. Bias Field Correction and Intensity Normalisation for Quantitative Analysis of Apparent Fibre Density. Paper presented at: International Society for Magnetic Resonance in Medicine 2017.

82. Royston P. Algorithm AS 181: The W test for Normality. Applied Statistics. 1982;31:176–180.

83. Royston P. Remark AS R94: A remark on Algorithm AS 181: The W test for normality. Applied Statistics. 1995;44:547–551.

84. Royston P. An extension of Shapiro and Wilk’s W test for normality to large samples. Applied Statistics. 1982;31:115–124.

85. R: A language and environment for statistical computing [computer program]. Vienna, Austria: R Foundation for Statistical Computing; 2020.

86. Durlak JA. How to select, calculate, and interpret effect sizes. Journal of pediatric psychology. Oct 2009;34(9):917–928.

87. Gibson B. Stretching for People with MS: An Illustrated Manual. In: Society NMS, ed. National Multiple Sclerosis Society: National Multiple Sclerosis Society; 2016.

