## Supplementary material for "Brain connectivity and motor improvements after ballet intervention in multiple sclerosis: pilot": GPower Protocol Resting State Functional Connectivity

*[1] -- Friday, February 26, 2021 -- 13:58:35*

**F tests** - ANOVA: Repeated measures, within-between interaction

| | | | |
|---|---|---|---|
| **Analysis:** | A priori: Compute required sample size | | |
| **Input:** | Effect size f | = | 0.303 |
| | α err prob | = | 0.005 |
| | Power (1-β err prob) | = | 0.80 |
| | Number of groups | = | 2 |
| | Number of measurements | = | 4 |
| | Corr among rep measures | = | 0.72 |
| | Nonsphericity correction ε | = | 1 |
| **Output:** | Noncentrality parameter λ | = | 20.9849143 |
| | Critical F | = | 4.9396168 |
| | Numerator df | = | 3.0000000 |
| | Denominator df | = | 42.0000000 |
| | Total sample size | = | 16 |
| | Actual power | = | 0.8198112 |
