## Supplementary material for "Brain connectivity and motor improvements after ballet intervention in multiple sclerosis: pilot": GPower Protocol StructuralConnectivity

*[1] -- Friday, February 26, 2021 -- 13:56:32*

**F tests** - ANOVA: Repeated measures, within-between interaction

| | | | |
|---|---|---|---|
| **Analysis:** | A priori: Compute required sample size | | |
| **Input:** | Effect size f | = | 0.0855 |
| | $\alpha$ err prob | = | 0.005 |
| | Power (1−$\beta$ err prob) | = | 0.80 |
| | Number of groups | = | 2 |
| | Number of measurements | = | 4 |
| | Corr among rep measures | = | 0.72 |
| | Nonsphericity correction $\epsilon$ | = | 1 |
| **Output:** | Noncentrality parameter $\lambda$ | = | 17.7534643 |
| | Critical F | = | 4.3300168 |
| | Numerator df | = | 3.0000000 |
| | Denominator df | = | 504 |
| | Total sample size | = | 170 |
| | Actual power | = | 0.8063827 |
