## Supplementary material for "Brain connectivity and motor improvements after ballet intervention in multiple sclerosis: pilot": R Statistical Analysis Report

### TDP MS w MRI

Paul Camacho

February 25th, 2021

### Read in Data Table

```
library(readxl)
TDPMSwMRI2019RCAf <- read_excel("C:/users/pcama/Documents/MSMRI/clin.xlsx",col_names = TRUE, na
 = "NA")
View(TDPMSwMRI2019RCAf)

MBB_ses_A_pipeline_results <- read_excel("C:/users/pcama/Downloads/MBB/MBB/MBB_ses_A_pipeline_re
sults.xlsx",col_names = TRUE, na = "NA")
```

```
## New names:
## * id0 -> id0...51
## * id0 -> id0...63
## * id0 -> id0...80
## * id0 -> id0...101
```

```
MBB_ses.B_pipeline_results <- read_excel("C:/users/pcama/Downloads/MBB/MBB/MBB_ses-B_pipeline_re
sults.xlsx",col_names = TRUE, na = "NA")
```

```
## New names:
## * id0 -> id0...51
## * id0 -> id0...67
## * id0 -> id0...84
## * id0 -> id0...105
```

```
shapiro.test(TDPMSwMRI2019RCAf$ICARS_Score_Post-TDPMSwMRI2019RCAf$ICARS_Score_Pre)
```

```
##
##  Shapiro-Wilk normality test
##
## data:  TDPMSwMRI2019RCAf$ICARS_Score_Post - TDPMSwMRI2019RCAf$ICARS_Score_Pre
## W = 0.88482, p-value = 0.3317
```

```
boxplot(TDPMSwMRI2019RCAf$ICARS_Score_Post-TDPMSwMRI2019RCAf$ICARS_Score_Pre)
```

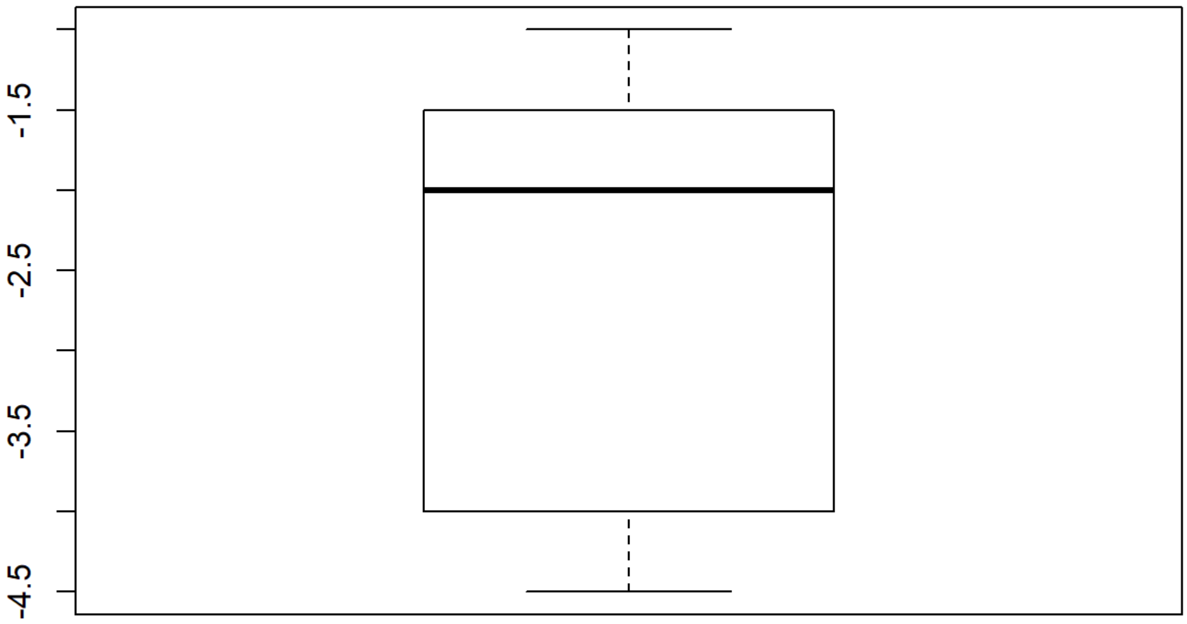


```
qqnorm(TDPMSwMRI2019RCAf$ICARS_Score_Post-TDPMSwMRI2019RCAf$ICARS_Score_Pre)
qqline(TDPMSwMRI2019RCAf$ICARS_Score_Post-TDPMSwMRI2019RCAf$ICARS_Score_Pre)
```

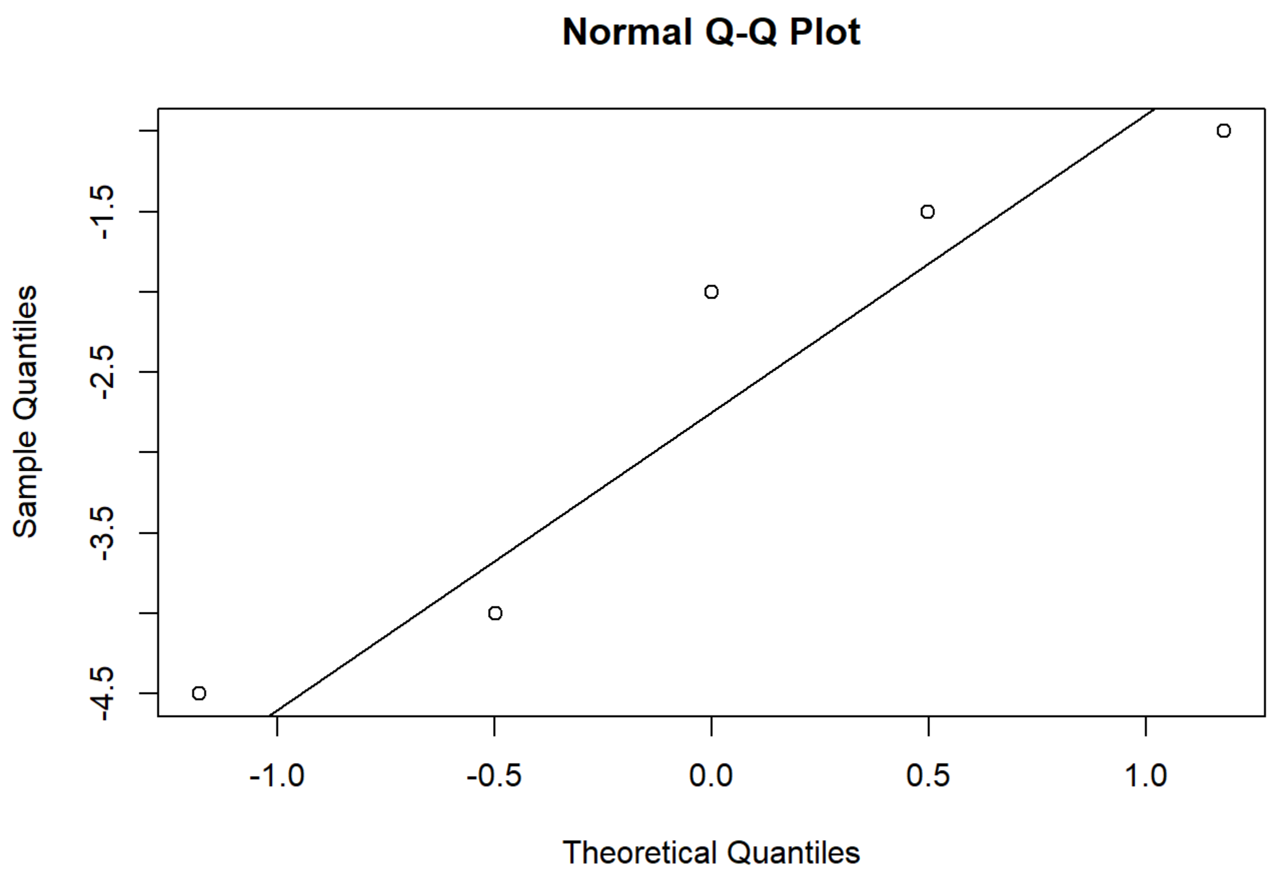


```
shapiro.test(TDPMSwMRI2019RCAf$ICARS_Posture_Gait_Post-TDPMSwMRI2019RCAf$ICARS_Posture_Gait_Pre)
```

```
##
##  Shapiro-Wilk normality test
##
## data:  TDPMSwMRI2019RCAf$ICARS_Posture_Gait_Post - TDPMSwMRI2019RCAf$ICARS_Posture_Gait_Pre
## W = 0.89095, p-value = 0.3619
```

```
boxplot(TDPMSwMRI2019RCAf$ICARS_Posture_Gait_Post-TDPMSwMRI2019RCAf$ICARS_Posture_Gait_Pre)
```

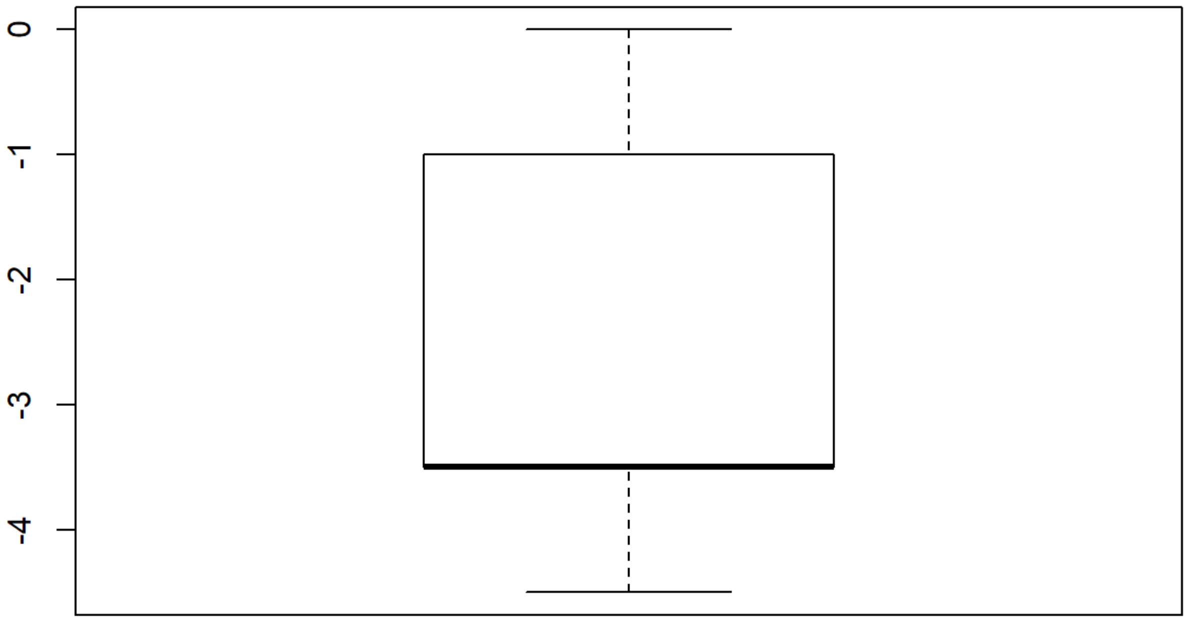


```
qqnorm(TDPMSwMRI2019RCAf$ICARS_Posture_Gait_Post-TDPMSwMRI2019RCAf$ICARS_Posture_Gait_Pre)
qqline(TDPMSwMRI2019RCAf$ICARS_Posture_Gait_Post-TDPMSwMRI2019RCAf$ICARS_Posture_Gait_Pre)
```

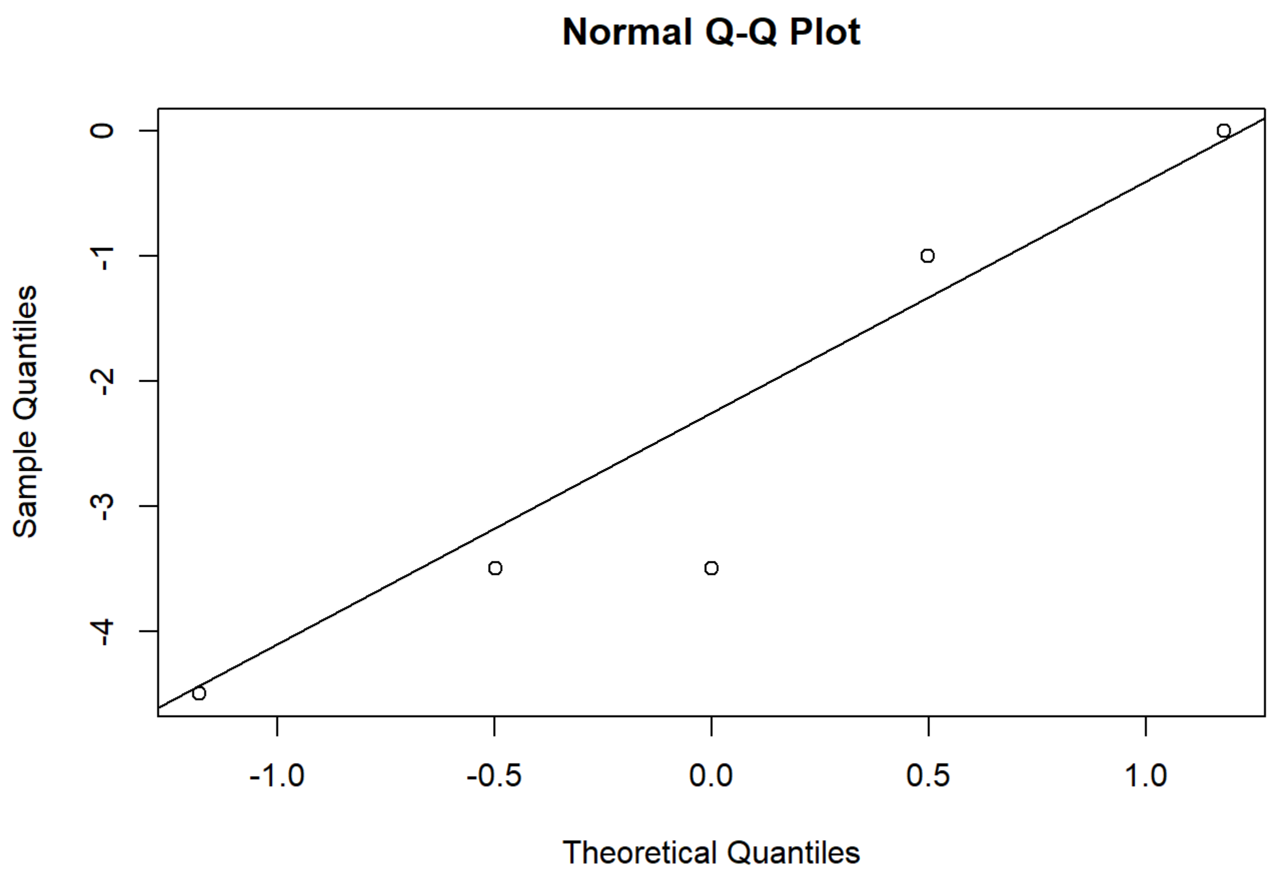


```
shapiro.test(TDPMSwMRI2019RCAf$ICARS_Kinetic_Function_Post-TDPMSwMRI2019RCAf$ICARS_Kinetic_Funct
ion_Pre)
```

```
##
##  Shapiro-Wilk normality test
##
## data:  TDPMSwMRI2019RCAf$ICARS_Kinetic_Function_Post - TDPMSwMRI2019RCAf$ICARS_Kinetic_Functi
on_Pre
## W = 0.80588, p-value = 0.09041
```

```
boxplot(TDPMSwMRI2019RCAf$ICARS_Kinetic_Function_Post-TDPMSwMRI2019RCAf$ICARS_Kinetic_Function_P
re)
```

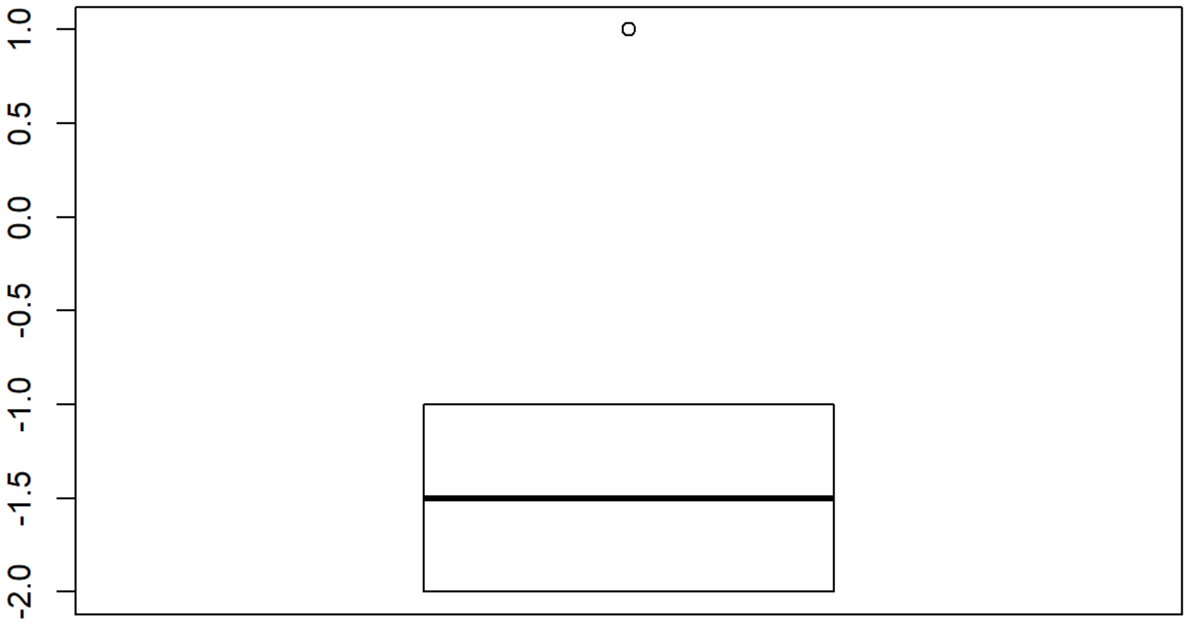


```
qqnorm(TDPMSwMRI2019RCAf$ICARS_Kinetic_Function_Post-TDPMSwMRI2019RCAf$ICARS_Kinetic_Function_Pr
e)
qqline(TDPMSwMRI2019RCAf$ICARS_Kinetic_Function_Post-TDPMSwMRI2019RCAf$ICARS_Kinetic_Function_Pr
e)
```

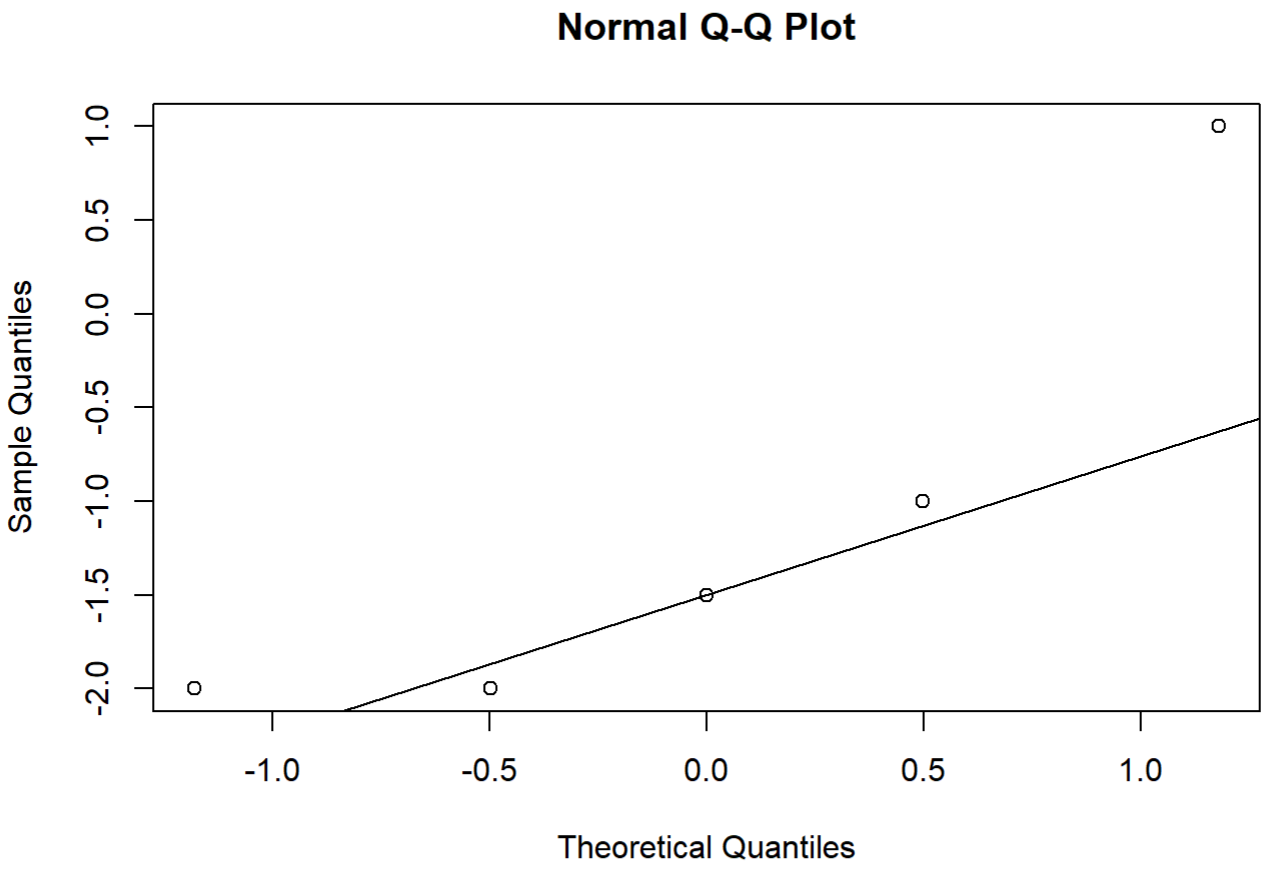


```
shapiro.test(TDPMSwMRI2019RCAf$Mini_BESTest_Score_Post-TDPMSwMRI2019RCAf$Mini_BESTest_Score_Pre)
```

```
##
##  Shapiro-Wilk normality test
##
## data:  TDPMSwMRI2019RCAf$Mini_BESTest_Score_Post - TDPMSwMRI2019RCAf$Mini_BESTest_Score_Pre
## W = 0.95563, p-value = 0.7773
```

```
boxplot(TDPMSwMRI2019RCAf$Mini_BESTest_Score_Post-TDPMSwMRI2019RCAf$Mini_BESTest_Score_Pre)
```

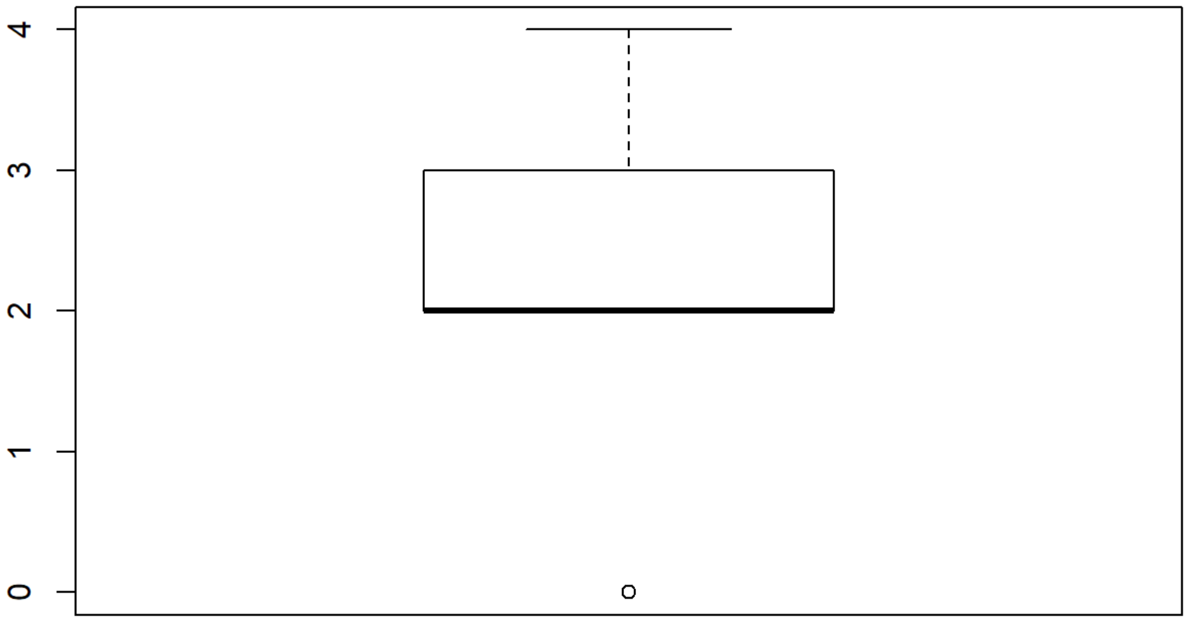


```
qqnorm(TDPMSwMRI2019RCAf$Mini_BESTest_Score_Post-TDPMSwMRI2019RCAf$Mini_BESTest_Score_Pre)
qqline(TDPMSwMRI2019RCAf$Mini_BESTest_Score_Post-TDPMSwMRI2019RCAf$Mini_BESTest_Score_Pre)
```

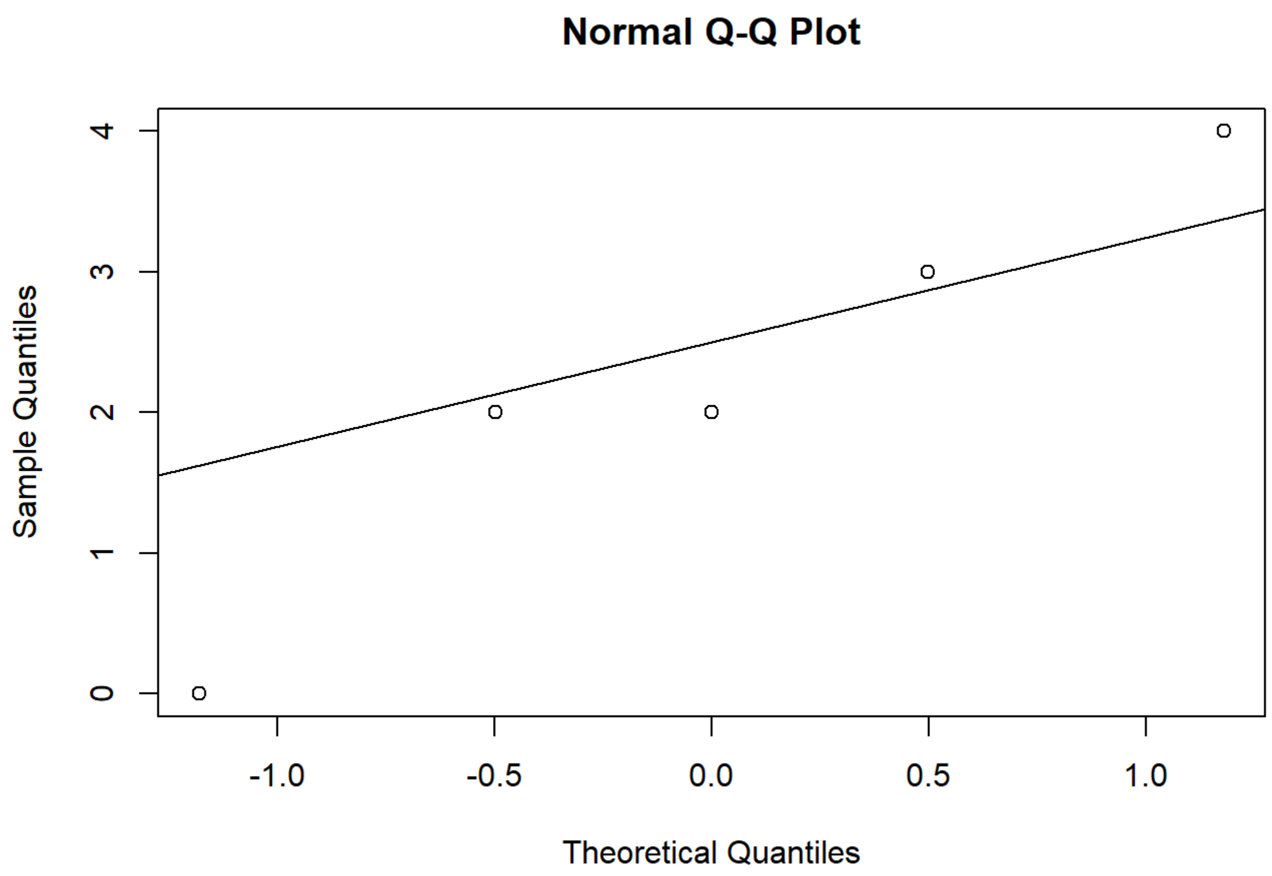


```
shapiro.test(TDPMSwMRI2019RCAf$s_index_post-TDPMSwMRI2019RCAf$s_index_pre)
```

```
##
##  Shapiro-Wilk normality test
##
## data:  TDPMSwMRI2019RCAf$s_index_post - TDPMSwMRI2019RCAf$s_index_pre
## W = 0.98038, p-value = 0.9366
```

```
boxplot(TDPMSwMRI2019RCAf$s_index_post-TDPMSwMRI2019RCAf$s_index_pre)
```

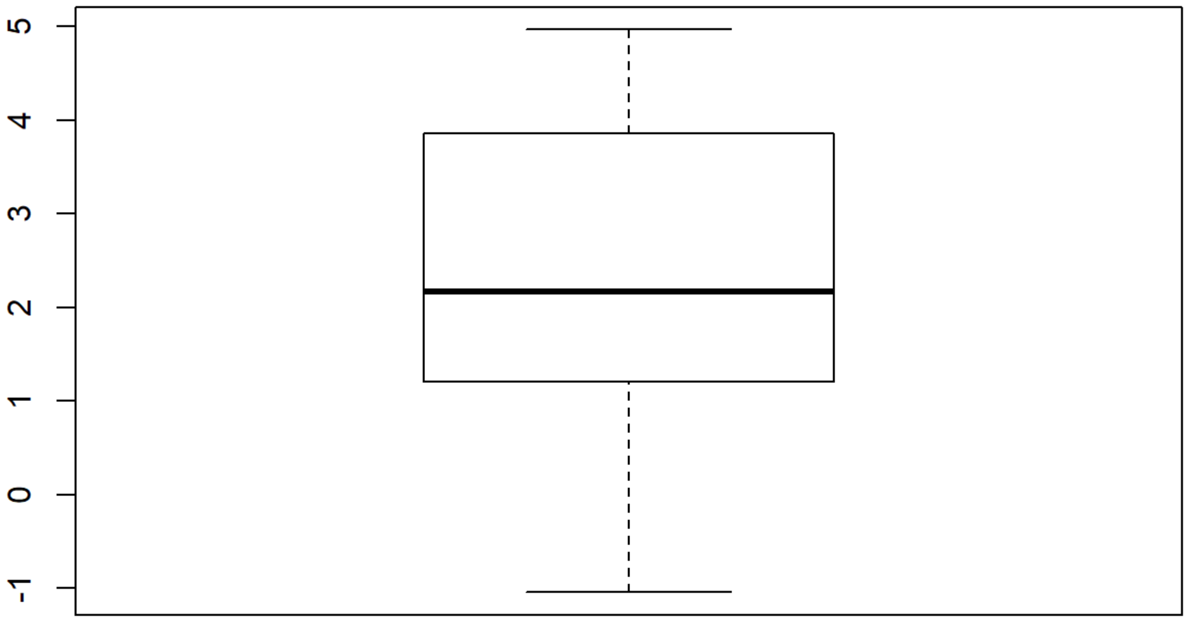


```
qqnorm(TDPMSwMRI2019RCAf$s_index_post-TDPMSwMRI2019RCAf$s_index_pre)
qqline(TDPMSwMRI2019RCAf$s_index_post-TDPMSwMRI2019RCAf$s_index_pre)
```

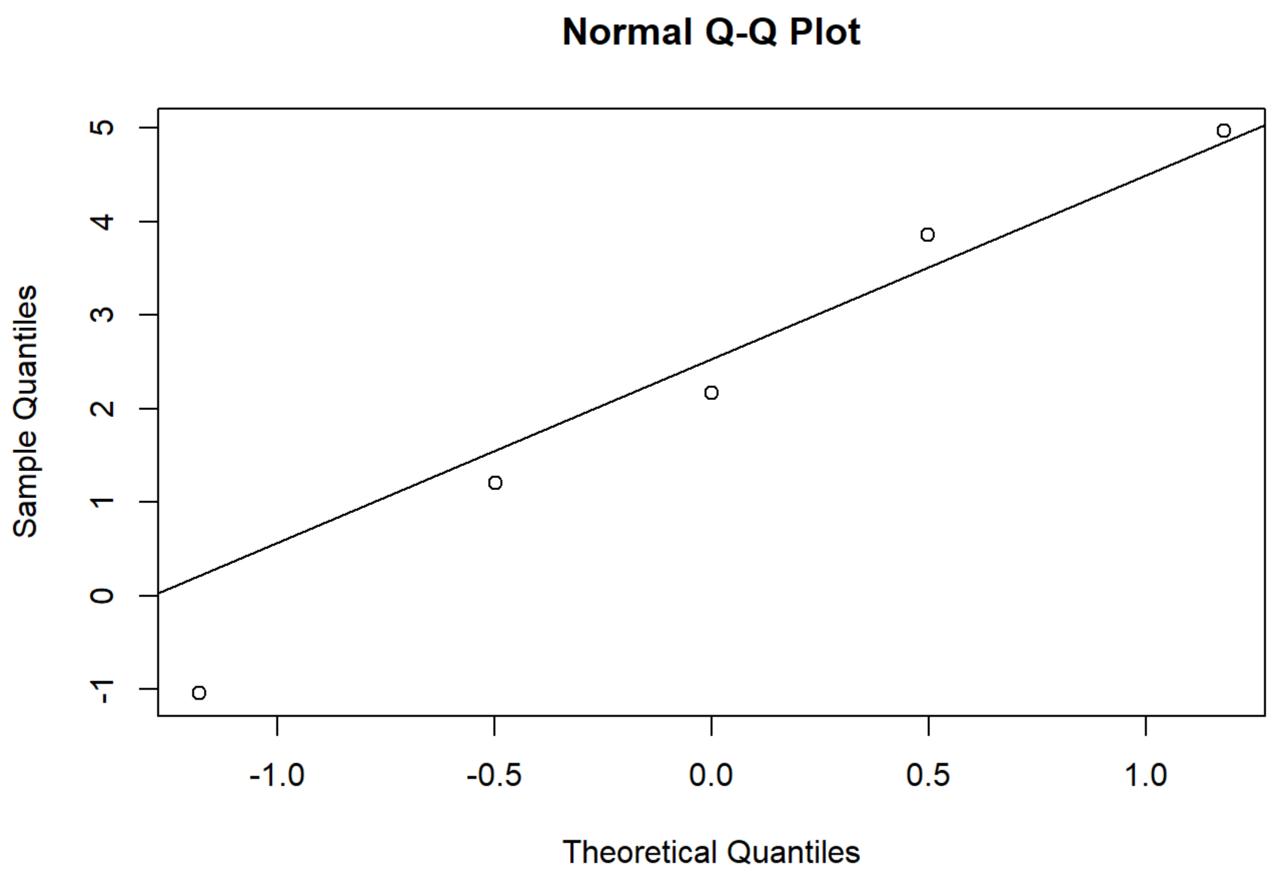


```
shapiro.test(MBB_ses.B_pipeline_results$GlobalEfficiencyAAL116SC-MBB_ses_A_pipeline_results$Glob
alEfficiencyAAL116SC)
```

```
##
##  Shapiro-Wilk normality test
##
## data:  MBB_ses.B_pipeline_results$GlobalEfficiencyAAL116SC - MBB_ses_A_pipeline_results$Globa
lEfficiencyAAL116SC
## W = 0.92837, p-value = 0.5853
```

```
boxplot(MBB_ses.B_pipeline_results$GlobalEfficiencyAAL116SC-MBB_ses_A_pipeline_results$GlobalEff
iciencyAAL116SC)
```

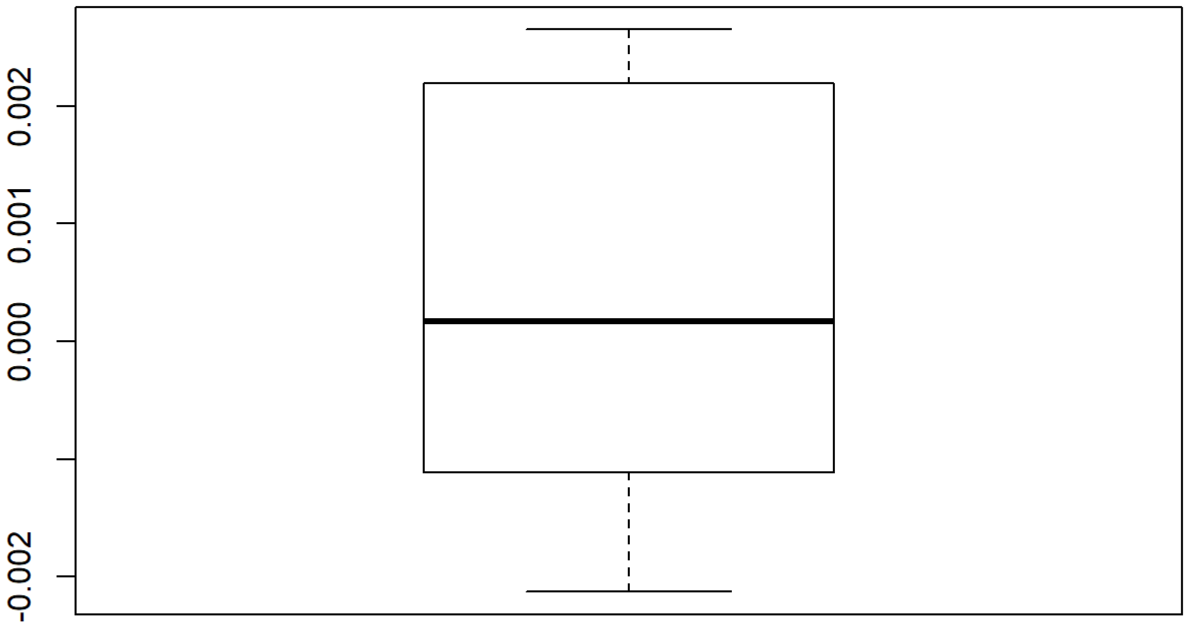


```
qqnorm(MBB_ses.B_pipeline_results$GlobalEfficiencyAAL116SC-MBB_ses_A_pipeline_results$GlobalEfficiencyAAL116SC)
qqline(MBB_ses.B_pipeline_results$GlobalEfficiencyAAL116SC-MBB_ses_A_pipeline_results$GlobalEfficiencyAAL116SC)
```

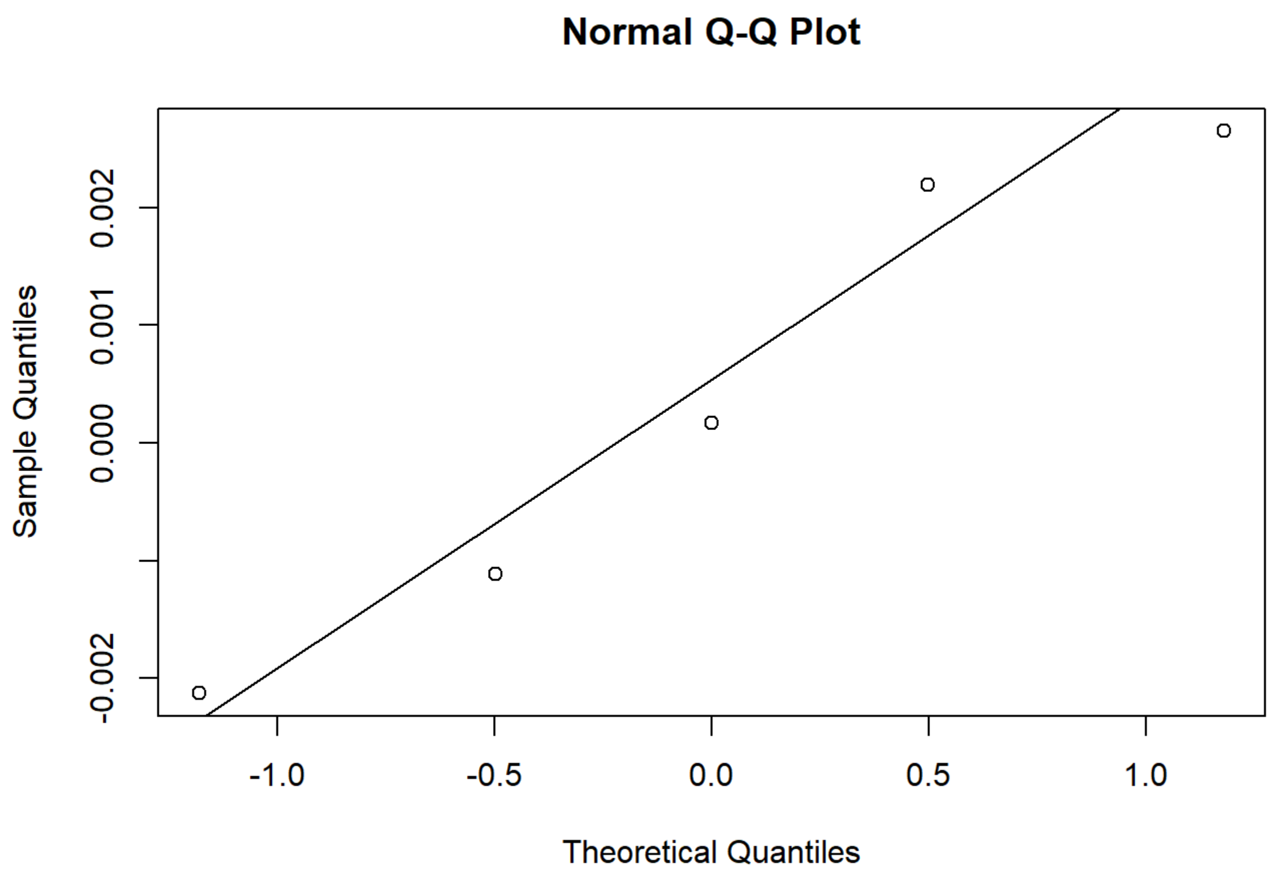


```
shapiro.test(MBB_ses.B_pipeline_results$MeanStrengthAAL116SC-MBB_ses_A_pipeline_results$MeanStre
ngthAAL116SC)
```

```
##
##  Shapiro-Wilk normality test
##
## data:  MBB_ses.B_pipeline_results$MeanStrengthAAL116SC - MBB_ses_A_pipeline_results$MeanStren
gthAAL116SC
## W = 0.87277, p-value = 0.2778
```

```
boxplot(MBB_ses.B_pipeline_results$MeanStrengthAAL116SC-MBB_ses_A_pipeline_results$MeanStrengthA
AL116SC)
```

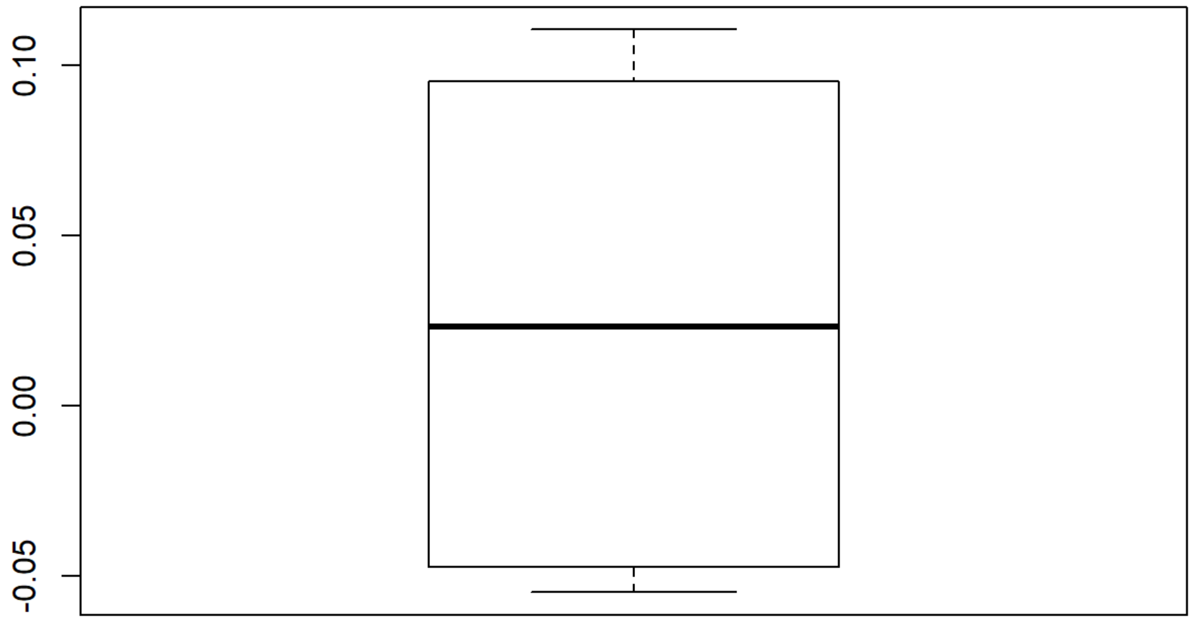


```
qqnorm(MBB_ses.B_pipeline_results$MeanStrengthAAL116SC-MBB_ses_A_pipeline_results$MeanStrengthAAL116SC)
qqline(MBB_ses.B_pipeline_results$MeanStrengthAAL116SC-MBB_ses_A_pipeline_results$MeanStrengthAAL116SC)
```

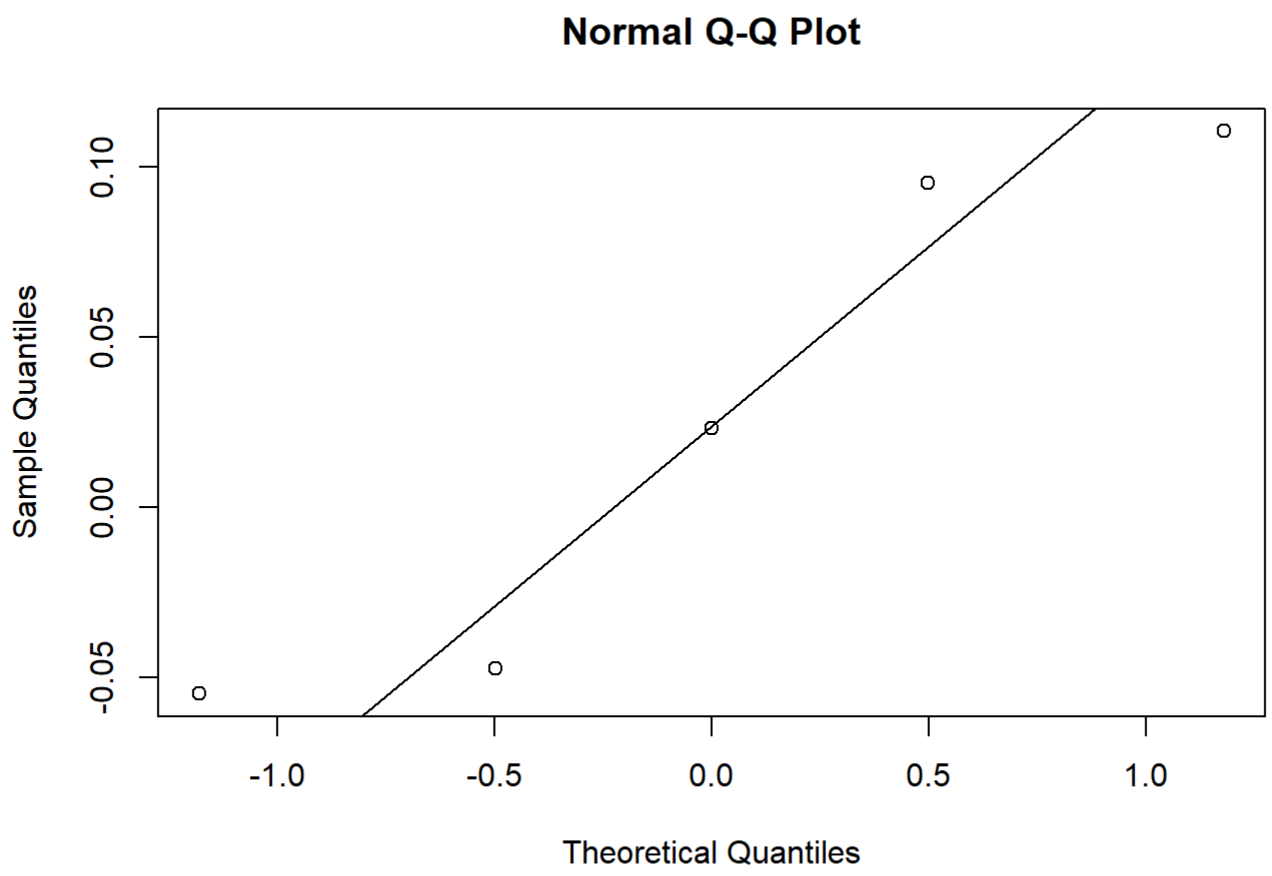


```
shapiro.test(MBB_ses.B_pipeline_results$MeanClusteringCoeffAAL116SC-MBB_ses_A_pipeline_results$MeanClusteringCoeffAAL116SC)
```

```
##
##  Shapiro-Wilk normality test
##
## data:  MBB_ses.B_pipeline_results$MeanClusteringCoeffAAL116SC - MBB_ses_A_pipeline_results$MeanClusteringCoeffAAL116SC
## W = 0.87787, p-value = 0.2998
```

```
boxplot(MBB_ses.B_pipeline_results$MeanClusteringCoeffAAL116SC-MBB_ses_A_pipeline_results$MeanClusteringCoeffAAL116SC)
```

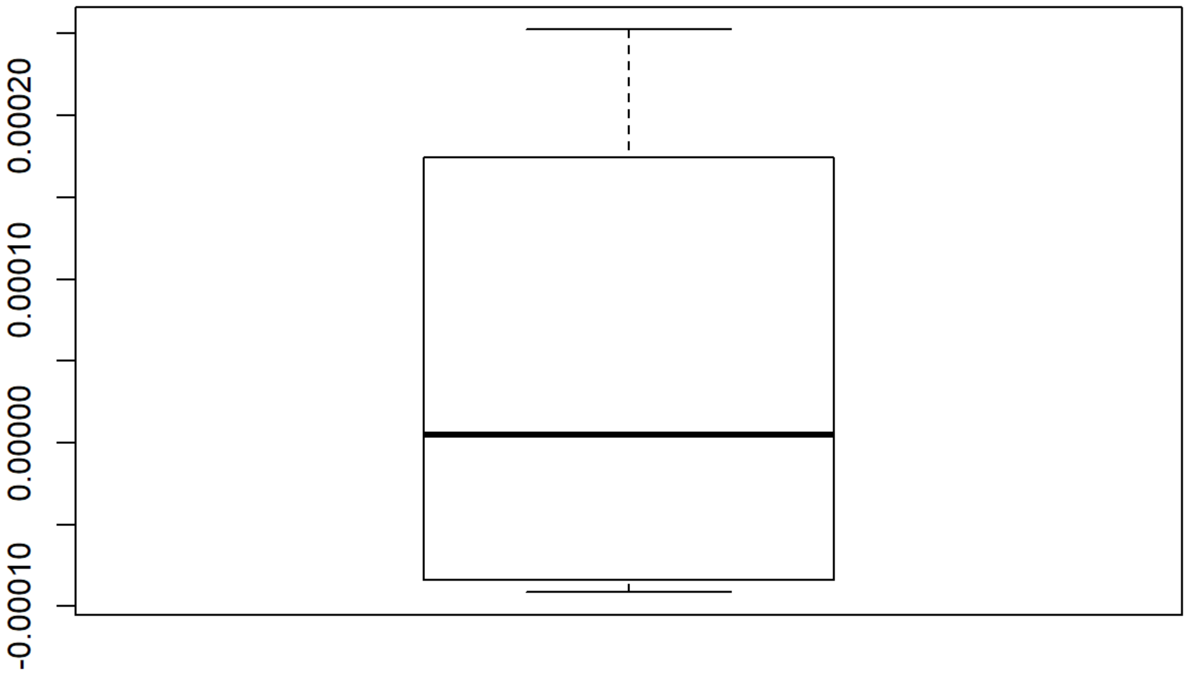


```
qqnorm(MBB_ses.B_pipeline_results$MeanClusteringCoeffAAL116SC-MBB_ses_A_pipeline_results$MeanClusteringCoeffAAL116SC)
qqline(MBB_ses.B_pipeline_results$MeanClusteringCoeffAAL116SC-MBB_ses_A_pipeline_results$MeanClusteringCoeffAAL116SC)
```

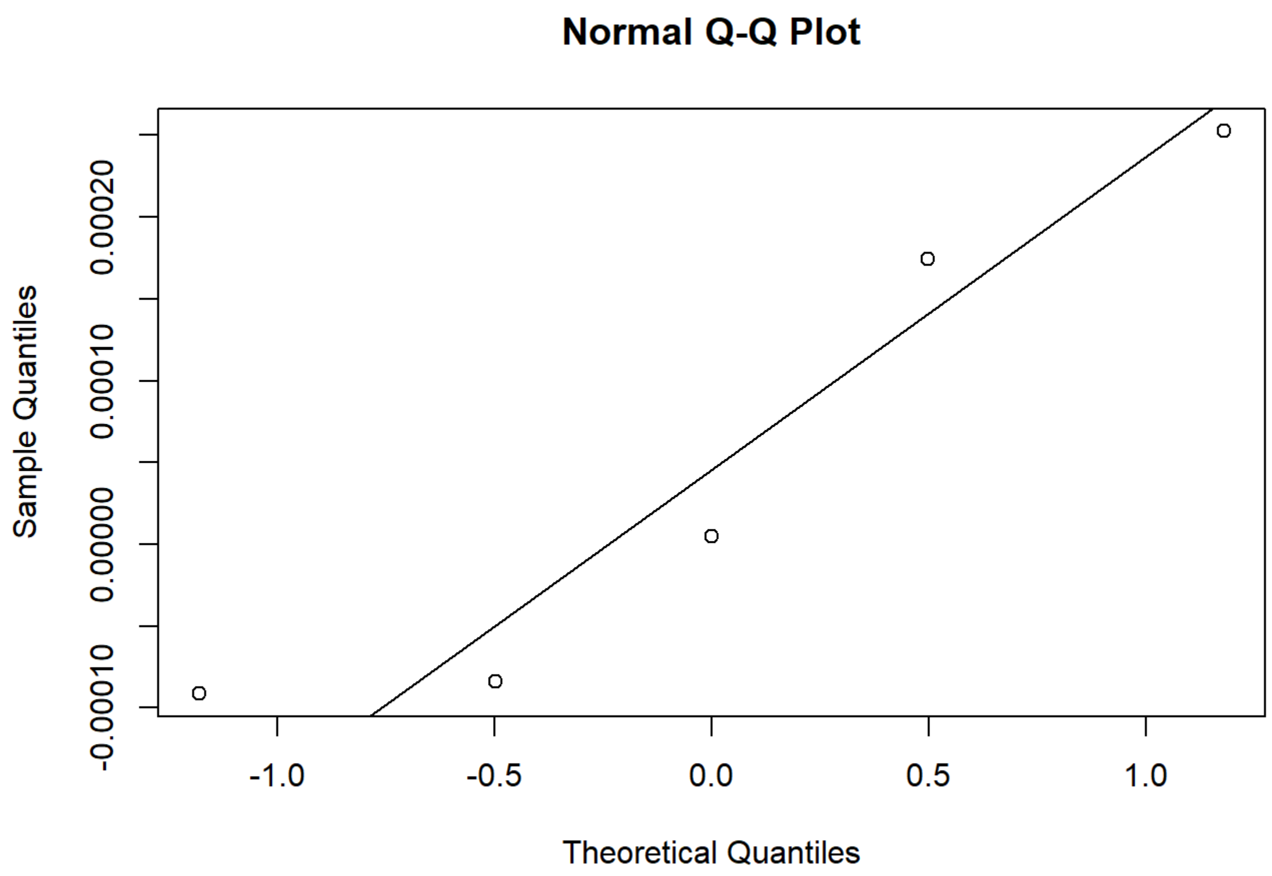


```
shapiro.test(MBB_ses.B_pipeline_results$GlobalEfficiencyaal116despike-MBB_ses_A_pipeline_results
$GlobalEfficiencyaal116despike)
```

```
##
##  Shapiro-Wilk normality test
##
## data:  MBB_ses.B_pipeline_results$GlobalEfficiencyaal116despike - MBB_ses_A_pipeline_results
$GlobalEfficiencyaal116despike
## W = 0.9763, p-value = 0.9139
```

```
boxplot(MBB_ses.B_pipeline_results$GlobalEfficiencyaal116despike-MBB_ses_A_pipeline_results$Glob
alEfficiencyaal116despike)
```

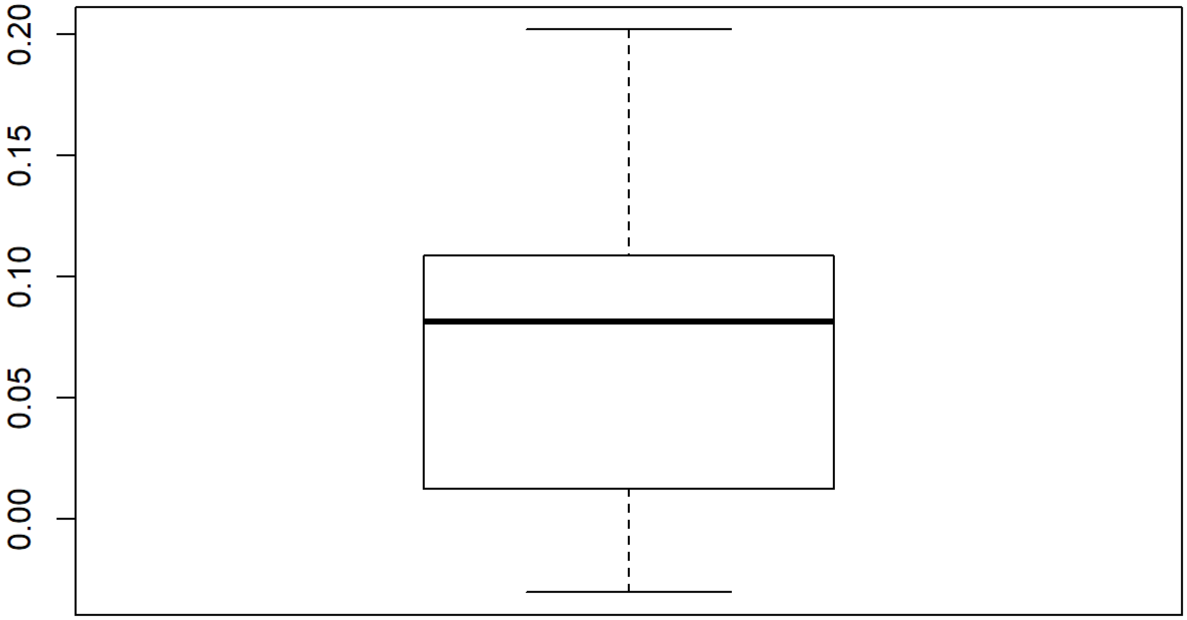


```
qqnorm(MBB_ses.B_pipeline_results$GlobalEfficiencyaal116despike-MBB_ses_A_pipeline_results$Globa
lEfficiencyaal116despike)
qqline(MBB_ses.B_pipeline_results$GlobalEfficiencyaal116despike-MBB_ses_A_pipeline_results$Globa
lEfficiencyaal116despike)
```

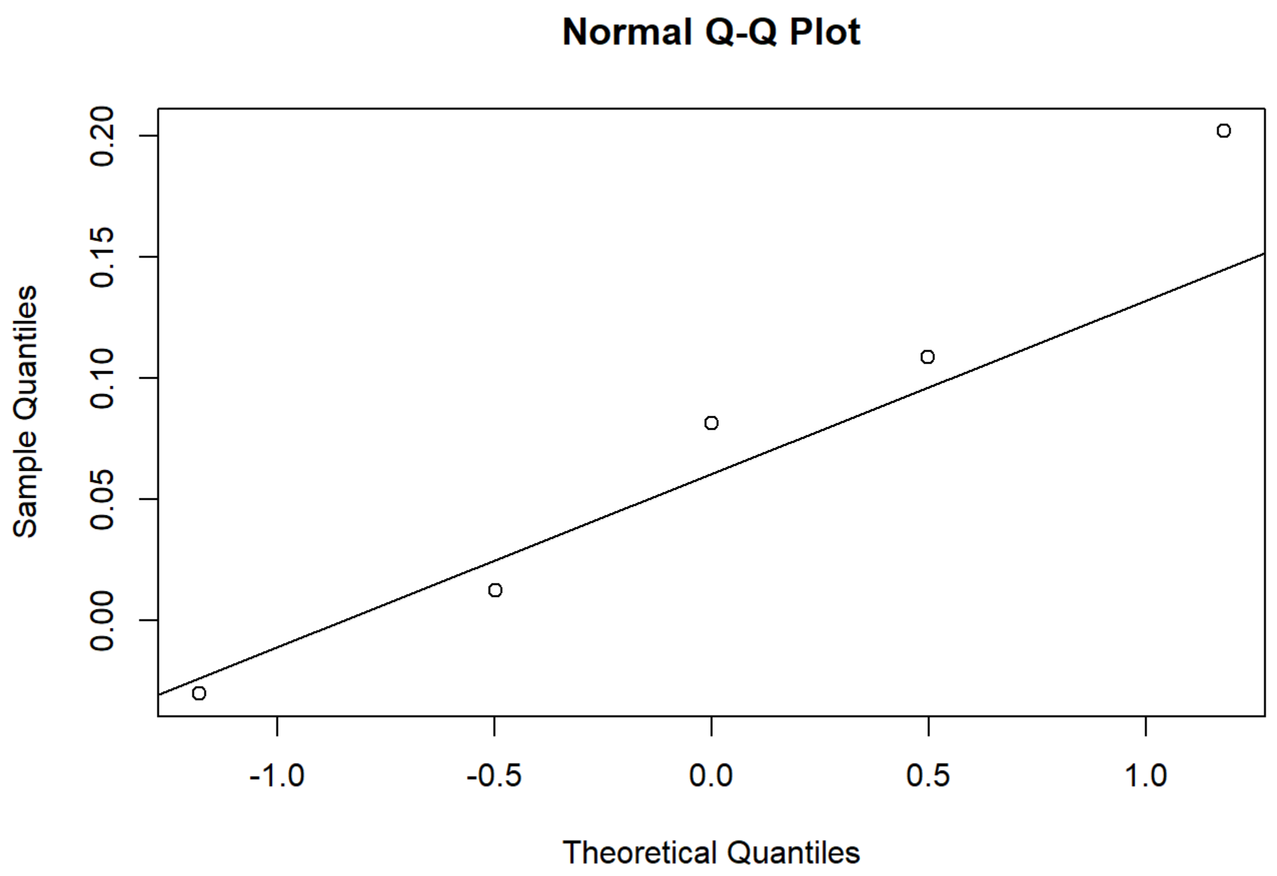


```
shapiro.test(MBB_ses.B_pipeline_results$MeanStrengthaal116despike-MBB_ses_A_pipeline_results$Mea
nStrengthaal116despike)
```

```
##
##  Shapiro-Wilk normality test
##
## data:  MBB_ses.B_pipeline_results$MeanStrengthaal116despike - MBB_ses_A_pipeline_results$Mean
Strengthaal116despike
## W = 0.99148, p-value = 0.9846
```

```
boxplot(MBB_ses.B_pipeline_results$MeanStrengthaal116despike-MBB_ses_A_pipeline_results$MeanStre
ngthaal116despike)
```

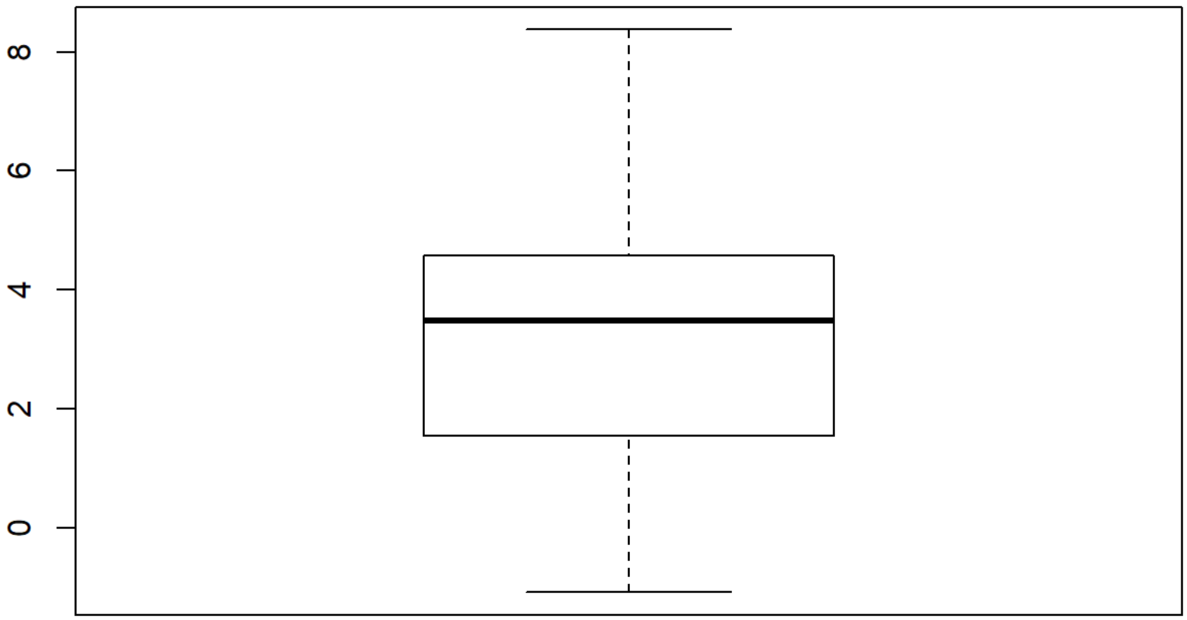


```
qqnorm(MBB_ses.B_pipeline_results$MeanStrengthaal116despike-MBB_ses_A_pipeline_results$MeanStren
gthaal116despike)
qqline(MBB_ses.B_pipeline_results$MeanStrengthaal116despike-MBB_ses_A_pipeline_results$MeanStren
gthaal116despike)
```

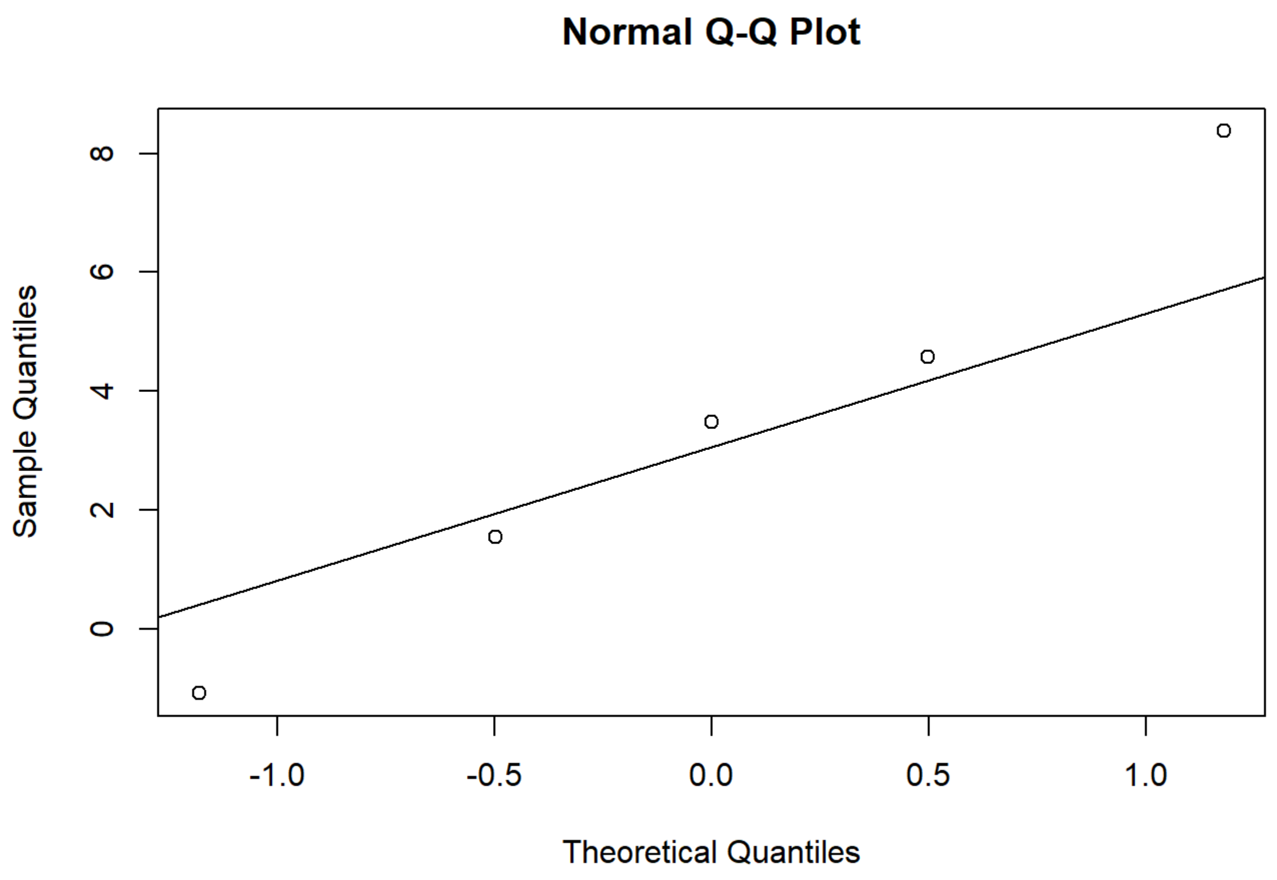


```
shapiro.test(MBB_ses.B_pipeline_results$MeanClusteringCoeffPosaal116despike-MBB_ses_A_pipeline_r
esults$MeanClusteringCoeffPosaal116despike)
```

```
##
##  Shapiro-Wilk normality test
##
## data:  MBB_ses.B_pipeline_results$MeanClusteringCoeffPosaal116despike -     MBB_ses_A_pipelin
e_results$MeanClusteringCoeffPosaal116despike
## W = 0.90425, p-value = 0.4338
```

```
boxplot(MBB_ses.B_pipeline_results$MeanClusteringCoeffPosaal116despike-MBB_ses_A_pipeline_result
s$MeanClusteringCoeffPosaal116despike)
```

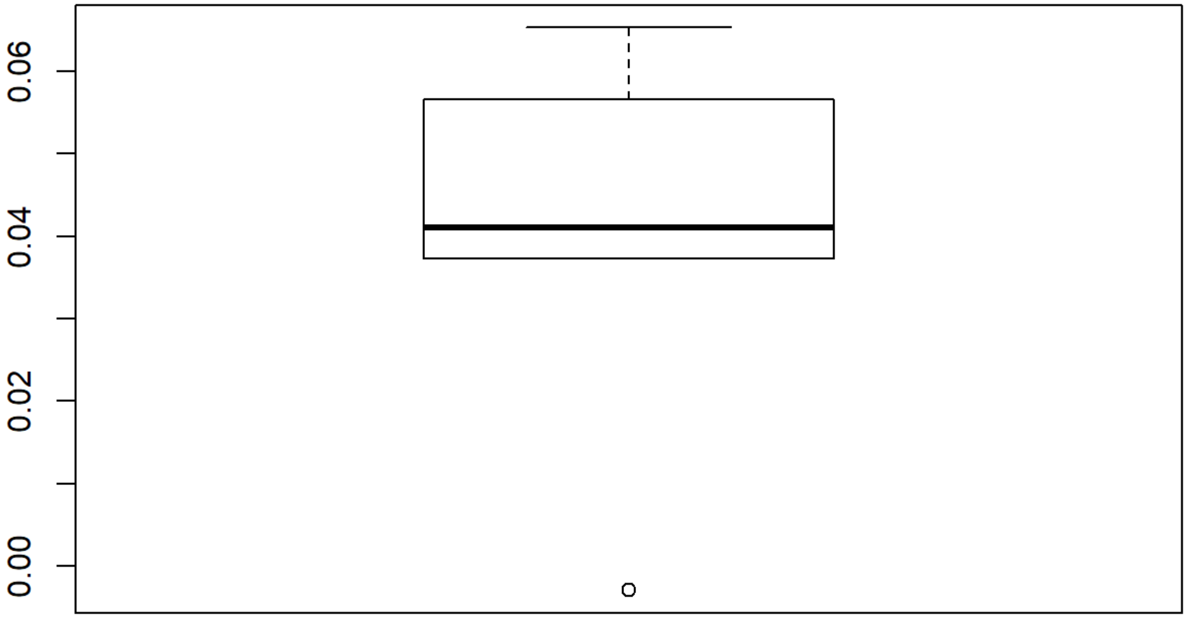


```
qqnorm(MBB_ses.B_pipeline_results$MeanClusteringCoeffPosaal116despike-MBB_ses_A_pipeline_results
$MeanClusteringCoeffPosaal116despike)
qqline(MBB_ses.B_pipeline_results$MeanClusteringCoeffPosaal116despike-MBB_ses_A_pipeline_results
$MeanClusteringCoeffPosaal116despike)
```

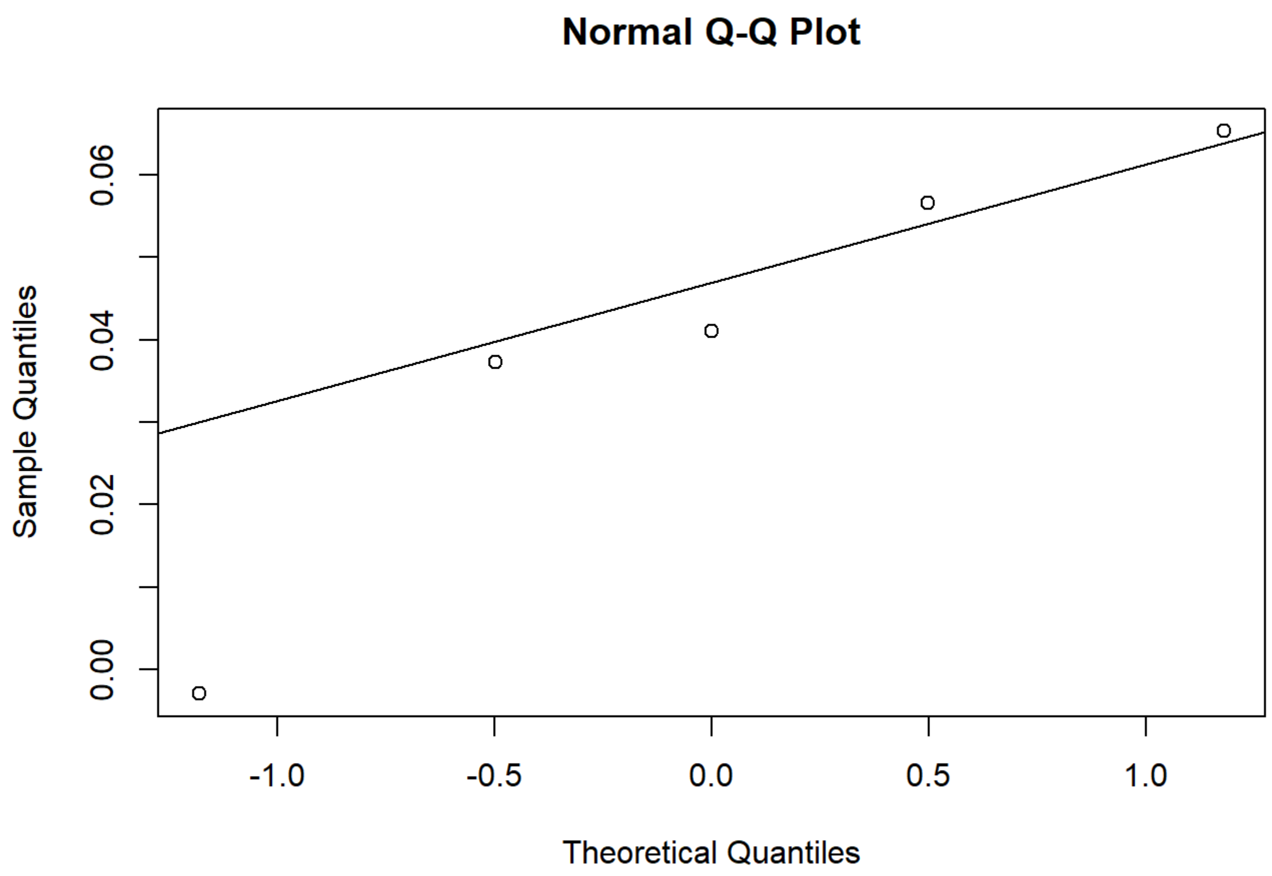


```
#t tests

t.test(TDPMSwMRI2019RCAf$ICARS_Score_Pre,TDPMSwMRI2019RCAf$ICARS_Score_Post, paired = TRUE, alte
rnative = "greater")
```

```
##
##  Paired t-test
##
## data:  TDPMSwMRI2019RCAf$ICARS_Score_Pre and TDPMSwMRI2019RCAf$ICARS_Score_Post
## t = 3.7334, df = 4, p-value = 0.01012
## alternative hypothesis: true difference in means is greater than 0
## 95 percent confidence interval:
##  1.115341      Inf
## sample estimates:
## mean of the differences
##                     2.6
```

```
t.test(TDPMSwMRI2019RCAf$ICARS_Posture_Gait_Pre,TDPMSwMRI2019RCAf$ICARS_Posture_Gait_Post, paire
d = TRUE, alternative = "greater")
```

```
##
##  Paired t-test
##
## data:  TDPMSwMRI2019RCAf$ICARS_Posture_Gait_Pre and TDPMSwMRI2019RCAf$ICARS_Posture_Gait_Post
## t = 2.9361, df = 4, p-value = 0.02128
## alternative hypothesis: true difference in means is greater than 0
## 95 percent confidence interval:
##  0.6847979       Inf
## sample estimates:
## mean of the differences
##                     2.5
```

```
t.test(TDPMSwMRI2019RCAf$ICARS_Kinetic_Function_Pre,TDPMSwMRI2019RCAf$ICARS_Kinetic_Function_Pos
t, paired = TRUE, alternative = "greater")
```

```
##
##  Paired t-test
##
## data:  TDPMSwMRI2019RCAf$ICARS_Kinetic_Function_Pre and TDPMSwMRI2019RCAf$ICARS_Kinetic_Funct
ion_Post
## t = 1.9757, df = 4, p-value = 0.0597
## alternative hypothesis: true difference in means is greater than 0
## 95 percent confidence interval:
##  -0.08696206         Inf
## sample estimates:
## mean of the differences
##                     1.1
```

```
t.test(TDPMSwMRI2019RCAf$Mini_BESTest_Score_Pre,TDPMSwMRI2019RCAf$Mini_BESTest_Score_Post, paire
d = TRUE, alternative = "less")
```

```
##
##  Paired t-test
##
## data:  TDPMSwMRI2019RCAf$Mini_BESTest_Score_Pre and TDPMSwMRI2019RCAf$Mini_BESTest_Score_Post
## t = -3.3166, df = 4, p-value = 0.01474
## alternative hypothesis: true difference in means is less than 0
## 95 percent confidence interval:
##        -Inf -0.7858928
## sample estimates:
## mean of the differences
##                    -2.2
```

```
t.test(TDPMSwMRI2019RCAf$s_index_pre,TDPMSwMRI2019RCAf$s_index_post, paired = TRUE, alternative
 = "less")
```

```
##
##  Paired t-test
##
## data:  TDPMSwMRI2019RCAf$s_index_pre and TDPMSwMRI2019RCAf$s_index_post
## t = -2.1328, df = 4, p-value = 0.04995
## alternative hypothesis: true difference in means is less than 0
## 95 percent confidence interval:
##           -Inf -0.0009648368
## sample estimates:
## mean of the differences
##                 -2.2334
```

```
t.test(MBB_ses.B_pipeline_results$GlobalEfficiencyaal116despike,MBB_ses_A_pipeline_results$Globa
lEfficiencyaal116despike, paired = TRUE, alternative = "greater")
```

```
##
##  Paired t-test
##
## data:  MBB_ses.B_pipeline_results$GlobalEfficiencyaal116despike and MBB_ses_A_pipeline_result
s$GlobalEfficiencyaal116despike
## t = 1.8655, df = 4, p-value = 0.06777
## alternative hypothesis: true difference in means is greater than 0
## 95 percent confidence interval:
##  -0.01068822         Inf
## sample estimates:
## mean of the differences
##              0.07485297
```

```
t.test(MBB_ses.B_pipeline_results$MeanStrengthaal116despike,MBB_ses_A_pipeline_results$MeanStren
gthaal116despike, paired = TRUE, alternative = "greater")
```

```
##
##  Paired t-test
##
## data:  MBB_ses.B_pipeline_results$MeanStrengthaal116despike and MBB_ses_A_pipeline_results$Me
anStrengthaal116despike
## t = 2.1479, df = 4, p-value = 0.04911
## alternative hypothesis: true difference in means is greater than 0
## 95 percent confidence interval:
##  0.02526466        Inf
## sample estimates:
## mean of the differences
##                3.382959
```

```
t.test(MBB_ses.B_pipeline_results$MeanClusteringCoeffPosaal116despike,MBB_ses_A_pipeline_results
$MeanClusteringCoeffPosaal116despike, paired = TRUE, alternative = "greater")
```

```
##
##  Paired t-test
##
## data:  MBB_ses.B_pipeline_results$MeanClusteringCoeffPosaal116despike and MBB_ses_A_pipeline_
results$MeanClusteringCoeffPosaal116despike
## t = 3.3531, df = 4, p-value = 0.01424
## alternative hypothesis: true difference in means is greater than 0
## 95 percent confidence interval:
##  0.01436504        Inf
## sample estimates:
## mean of the differences
##              0.03944075
```

```
t.test(MBB_ses.B_pipeline_results$GlobalEfficiencyAAL116SC,MBB_ses_A_pipeline_results$GlobalEffi
ciencyAAL116SC, paired = TRUE, alternative = "less")
```

```
##
##  Paired t-test
##
## data:  MBB_ses.B_pipeline_results$GlobalEfficiencyAAL116SC and MBB_ses_A_pipeline_results$Glo
balEfficiencyAAL116SC
## t = 0.38371, df = 4, p-value = 0.6396
## alternative hypothesis: true difference in means is less than 0
## 95 percent confidence interval:
##         -Inf 0.002319737
## sample estimates:
## mean of the differences
##             0.000353844
```

```
t.test(MBB_ses.B_pipeline_results$MeanClusteringCoeffAAL116SC,MBB_ses_A_pipeline_results$MeanClu
steringCoeffAAL116SC, paired = TRUE, alternative = "less")
```

```
##
##  Paired t-test
##
## data:  MBB_ses.B_pipeline_results$MeanClusteringCoeffAAL116SC and MBB_ses_A_pipeline_results
$MeanClusteringCoeffAAL116SC
## t = 0.74171, df = 4, p-value = 0.7503
## alternative hypothesis: true difference in means is less than 0
## 95 percent confidence interval:
##          -Inf 0.0001993786
## sample estimates:
## mean of the differences
##              5.1463e-05
```

```
t.test(MBB_ses.B_pipeline_results$MeanStrengthAAL116SC,MBB_ses_A_pipeline_results$MeanStrengthAA
L116SC, paired = TRUE, alternative = "less")
```

```
##
##  Paired t-test
##
## data:  MBB_ses.B_pipeline_results$MeanStrengthAAL116SC and MBB_ses_A_pipeline_results$MeanStr
engthAAL116SC
## t = 0.73714, df = 4, p-value = 0.749
## alternative hypothesis: true difference in means is less than 0
## 95 percent confidence interval:
##        -Inf 0.09900218
## sample estimates:
## mean of the differences
##              0.02543703
```

```
#nonparametric two-sided

wilcox.test(TDPMSwMRI2019RCAf$ICARS_Score_Pre,TDPMSwMRI2019RCAf$ICARS_Score_Post, paired = TRUE,
alternative = "greater")
```

```
##
##  Wilcoxon signed rank test
##
## data:  TDPMSwMRI2019RCAf$ICARS_Score_Pre and TDPMSwMRI2019RCAf$ICARS_Score_Post
## V = 15, p-value = 0.03125
## alternative hypothesis: true location shift is greater than 0
```

```
wilcox.test(TDPMSwMRI2019RCAf$ICARS_Posture_Gait_Pre,TDPMSwMRI2019RCAf$ICARS_Posture_Gait_Post,
paired = TRUE, alternative = "greater")
```

```
## Warning in wilcox.test.default(TDPMSwMRI2019RCAf$ICARS_Posture_Gait_Pre, :
## cannot compute exact p-value with ties
```

```
## Warning in wilcox.test.default(TDPMSwMRI2019RCAf$ICARS_Posture_Gait_Pre, :
## cannot compute exact p-value with zeroes
```

```
##
##  Wilcoxon signed rank test with continuity correction
##
## data:  TDPMSwMRI2019RCAf$ICARS_Posture_Gait_Pre and TDPMSwMRI2019RCAf$ICARS_Posture_Gait_Post
## V = 10, p-value = 0.04876
## alternative hypothesis: true location shift is greater than 0
```

```
wilcox.test(TDPMSwMRI2019RCAf$ICARS_Kinetic_Function_Pre,TDPMSwMRI2019RCAf$ICARS_Kinetic_Functio
n_Post, paired = TRUE, alternative = "greater")
```

```
## Warning in
## wilcox.test.default(TDPMSwMRI2019RCAf$ICARS_Kinetic_Function_Pre, : cannot
## compute exact p-value with ties
```

```
##
##  Wilcoxon signed rank test with continuity correction
##
## data:  TDPMSwMRI2019RCAf$ICARS_Kinetic_Function_Pre and TDPMSwMRI2019RCAf$ICARS_Kinetic_Funct
ion_Post
## V = 13.5, p-value = 0.06721
## alternative hypothesis: true location shift is greater than 0
```

```
wilcox.test(TDPMSwMRI2019RCAf$Mini_BESTest_Score_Pre,TDPMSwMRI2019RCAf$Mini_BESTest_Score_Post,
paired = TRUE, alternative = "less")
```

```
## Warning in wilcox.test.default(TDPMSwMRI2019RCAf$Mini_BESTest_Score_Pre, :
## cannot compute exact p-value with ties
```

```
## Warning in wilcox.test.default(TDPMSwMRI2019RCAf$Mini_BESTest_Score_Pre, :
## cannot compute exact p-value with zeroes
```

```
##
##  Wilcoxon signed rank test with continuity correction
##
## data:  TDPMSwMRI2019RCAf$Mini_BESTest_Score_Pre and TDPMSwMRI2019RCAf$Mini_BESTest_Score_Post
## V = 0, p-value = 0.04876
## alternative hypothesis: true location shift is less than 0
```

```
wilcox.test(TDPMSwMRI2019RCAf$s_index_pre,TDPMSwMRI2019RCAf$s_index_post, paired = TRUE, alterna
tive = "less")
```

```
##
##  Wilcoxon signed rank test
##
## data:  TDPMSwMRI2019RCAf$s_index_pre and TDPMSwMRI2019RCAf$s_index_post
## V = 1, p-value = 0.0625
## alternative hypothesis: true location shift is less than 0
```

```
wilcox.test(MBB_ses.B_pipeline_results$GlobalEfficiencyaal116despike,MBB_ses_A_pipeline_results
$GlobalEfficiencyaal116despike, paired = TRUE, alternative = "greater")
```

```
##
##  Wilcoxon signed rank test
##
## data:  MBB_ses.B_pipeline_results$GlobalEfficiencyaal116despike and MBB_ses_A_pipeline_result
s$GlobalEfficiencyaal116despike
## V = 13, p-value = 0.09375
## alternative hypothesis: true location shift is greater than 0
```

```
wilcox.test(MBB_ses.B_pipeline_results$MeanStrengthaal116despike,MBB_ses_A_pipeline_results$Mean
Strengthaal116despike, paired = TRUE, alternative = "less")
```

```
##
##  Wilcoxon signed rank test
##
## data:  MBB_ses.B_pipeline_results$MeanStrengthaal116despike and MBB_ses_A_pipeline_results$Me
anStrengthaal116despike
## V = 14, p-value = 0.9688
## alternative hypothesis: true location shift is less than 0
```

```
wilcox.test(MBB_ses.B_pipeline_results$MeanClusteringCoeffPosaal116despike,MBB_ses_A_pipeline_re
sults$MeanClusteringCoeffPosaal116despike, paired = TRUE, alternative = "less")
```

```
##
##  Wilcoxon signed rank test
##
## data:  MBB_ses.B_pipeline_results$MeanClusteringCoeffPosaal116despike and MBB_ses_A_pipeline_
results$MeanClusteringCoeffPosaal116despike
## V = 14, p-value = 0.9688
## alternative hypothesis: true location shift is less than 0
```

```
wilcox.test(MBB_ses.B_pipeline_results$GlobalEfficiencyAAL116SC,MBB_ses_A_pipeline_results$Globa
lEfficiencyAAL116SC, paired = TRUE, alternative = "less")
```

```
##
##  Wilcoxon signed rank test
##
## data:  MBB_ses.B_pipeline_results$GlobalEfficiencyAAL116SC and MBB_ses_A_pipeline_results$Glo
balEfficiencyAAL116SC
## V = 10, p-value = 0.7812
## alternative hypothesis: true location shift is less than 0
```

```
wilcox.test(MBB_ses.B_pipeline_results$MeanClusteringCoeffAAL116SC,MBB_ses_A_pipeline_results$Me
anClusteringCoeffAAL116SC, paired = TRUE, alternative = "less")
```

```
##
##  Wilcoxon signed rank test
##
## data:  MBB_ses.B_pipeline_results$MeanClusteringCoeffAAL116SC and MBB_ses_A_pipeline_results
$MeanClusteringCoeffAAL116SC
## V = 10, p-value = 0.7812
## alternative hypothesis: true location shift is less than 0
```

```
wilcox.test(MBB_ses.B_pipeline_results$MeanStrengthAAL116SC,MBB_ses_A_pipeline_results$MeanStren
gthAAL116SC, paired = TRUE, alternative = "less")
```

```
##
##  Wilcoxon signed rank test
##
## data:  MBB_ses.B_pipeline_results$MeanStrengthAAL116SC and MBB_ses_A_pipeline_results$MeanStr
engthAAL116SC
## V = 10, p-value = 0.7812
## alternative hypothesis: true location shift is less than 0
```

### Effect Sizes

```
library(effsize)
cohen.d(TDPMSwMRI2019RCAf$ICARS_Score_Post-TDPMSwMRI2019RCAf$ICARS_Score_Pre, f = NA, paired=TRU
E, within=TRUE, hedges.correction=FALSE,conf.level=0.95,pooled=TRUE)
```

```
##
## Cohen's d (single sample)
##
## d estimate: -1.669619 (large)
## Reference mu: 0
## 95 percent confidence interval:
##     lower     upper
## -4.553333  1.214094
```

```
cohen.d(TDPMSwMRI2019RCAf$ICARS_Score_Post-TDPMSwMRI2019RCAf$ICARS_Score_Pre, f = NA, paired=TRU
E, within=TRUE, hedges.correction=TRUE,conf.level=0.95,pooled=TRUE)
```

```
##
## Hedges's g (single sample)
##
## g estimate: -1.214269 (large)
## Reference mu: 0
## 95 percent confidence interval:
##      lower      upper
## -3.1797248  0.7511875
```

```
cohen.d(TDPMSwMRI2019RCAf$ICARS_Posture_Gait_Post-TDPMSwMRI2019RCAf$ICARS_Posture_Gait_Pre, f =
NA, paired=TRUE, within=TRUE, hedges.correction=FALSE,conf.level=0.95,pooled=TRUE)
```

```
##
## Cohen's d (single sample)
##
## d estimate: -1.313064 (large)
## Reference mu: 0
## 95 percent confidence interval:
##     lower     upper
## -4.050946  1.424817
```

```
cohen.d(TDPMSwMRI2019RCAf$ICARS_Posture_Gait_Post-TDPMSwMRI2019RCAf$ICARS_Posture_Gait_Pre, f =
NA, paired=TRUE, within=TRUE, hedges.correction=TRUE,conf.level=0.95,pooled=TRUE)
```

```
##
## Hedges's g (single sample)
##
## g estimate: -0.9549559 (large)
## Reference mu: 0
## 95 percent confidence interval:
##     lower     upper
## -2.861174  0.951262
```

```
cohen.d(TDPMSwMRI2019RCAf$Mini_BESTest_Score_Post-TDPMSwMRI2019RCAf$Mini_BESTest_Score_Pre, f =
NA, paired=TRUE, within=TRUE, hedges.correction=FALSE,conf.level=0.95,pooled=TRUE)
```

```
##
## Cohen's d (single sample)
##
## d estimate: 1.48324 (large)
## Reference mu: 0
## 95 percent confidence interval:
##     lower     upper
## -1.320832  4.287312
```

```
cohen.d(TDPMSwMRI2019RCAf$Mini_BESTest_Score_Post-TDPMSwMRI2019RCAf$Mini_BESTest_Score_Pre, f =
NA, paired=TRUE, within=TRUE, hedges.correction=TRUE,conf.level=0.95,pooled=TRUE)
```

```
##
## Hedges's g (single sample)
##
## g estimate: 1.07872 (large)
## Reference mu: 0
## 95 percent confidence interval:
##      lower      upper
## -0.8542289  3.0116684
```

```
cohen.d(TDPMSwMRI2019RCAf$s_index_post-TDPMSwMRI2019RCAf$s_index_pre, f = NA, paired=TRUE, withi
n=TRUE, hedges.correction=FALSE,conf.level=0.95,pooled=TRUE)
```

```
##
## Cohen's d (single sample)
##
## d estimate: 0.9538029 (large)
## Reference mu: 0
## 95 percent confidence interval:
##     lower     upper
## -1.666923  3.574529
```

```
cohen.d(TDPMSwMRI2019RCAf$s_index_post-TDPMSwMRI2019RCAf$s_index_pre, f = NA, paired=TRUE, withi
n=TRUE, hedges.correction=TRUE,conf.level=0.95,pooled=TRUE)
```

```
##
## Hedges's g (single sample)
##
## g estimate: 0.6936748 (medium)
## Reference mu: 0
## 95 percent confidence interval:
##     lower     upper
## -1.165904  2.553254
```

```
cohen.d(MBB_ses.B_pipeline_results$GlobalEfficiencyAAL116SC-MBB_ses_A_pipeline_results$GlobalEff
iciencyAAL116SC, f = NA, paired=TRUE, within=TRUE, hedges.correction=FALSE,conf.level=0.95,poole
d=TRUE)
```

```
##
## Cohen's d (single sample)
##
## d estimate: 0.1716023 (negligible)
## Reference mu: 0
## 95 percent confidence interval:
##     lower     upper
## -2.316292  2.659497
```

```
cohen.d(MBB_ses.B_pipeline_results$GlobalEfficiencyAAL116SC-MBB_ses_A_pipeline_results$GlobalEff
iciencyAAL116SC, f = NA, paired=TRUE, within=TRUE, hedges.correction=TRUE,conf.level=0.95,pooled
=TRUE)
```

```
##
## Hedges's g (single sample)
##
## g estimate: 0.1248016 (negligible)
## Reference mu: 0
## 95 percent confidence interval:
##     lower     upper
## -1.683012  1.932616
```

```
cohen.d(MBB_ses.B_pipeline_results$MeanStrengthAAL116SC-MBB_ses_A_pipeline_results$MeanStrengthA
AL116SC, f = NA, paired=TRUE, within=TRUE, hedges.correction=FALSE,conf.level=0.95,pooled=TRUE)
```

```
##
## Cohen's d (single sample)
##
## d estimate: 0.3296592 (small)
## Reference mu: 0
## 95 percent confidence interval:
##     lower     upper
## -2.170479  2.829798
```

```
cohen.d(MBB_ses.B_pipeline_results$MeanStrengthAAL116SC-MBB_ses_A_pipeline_results$MeanStrengthA
AL116SC, f = NA, paired=TRUE, within=TRUE, hedges.correction=TRUE,conf.level=0.95,pooled=TRUE)
```

```
##
## Hedges's g (single sample)
##
## g estimate: 0.2397521 (small)
## Reference mu: 0
## 95 percent confidence interval:
##     lower     upper
## -1.572781  2.052286
```

```
cohen.d(MBB_ses.B_pipeline_results$MeanClusteringCoeffAAL116SC-MBB_ses_A_pipeline_results$MeanCl
usteringCoeffAAL116SC, f = NA, paired=TRUE, within=TRUE, hedges.correction=FALSE,conf.level=0.95
,pooled=TRUE)
```

```
##
## Cohen's d (single sample)
##
## d estimate: 0.331705 (small)
## Reference mu: 0
## 95 percent confidence interval:
##     lower     upper
## -2.168642  2.832052
```

```
cohen.d(MBB_ses.B_pipeline_results$MeanClusteringCoeffAAL116SC-MBB_ses_A_pipeline_results$MeanCl
usteringCoeffAAL116SC, f = NA, paired=TRUE, within=TRUE, hedges.correction=TRUE,conf.level=0.95,
pooled=TRUE)
```

```
##
## Hedges's g (single sample)
##
## g estimate: 0.24124 (small)
## Reference mu: 0
## 95 percent confidence interval:
##     lower     upper
## -1.571374  2.053854
```

```
cohen.d(MBB_ses.B_pipeline_results$GlobalEfficiencyaal116despike-MBB_ses_A_pipeline_results$Glob
alEfficiencyaal116despike, f = NA, paired=TRUE, within=TRUE, hedges.correction=FALSE,conf.level=
0.95,pooled=TRUE)
```

```
##
## Cohen's d (single sample)
##
## d estimate: 0.8342664 (large)
## Reference mu: 0
## 95 percent confidence interval:
##     lower     upper
## -1.754834  3.423367
```

```
cohen.d(MBB_ses.B_pipeline_results$GlobalEfficiencyaal116despike-MBB_ses_A_pipeline_results$Glob
alEfficiencyaal116despike, f = NA, paired=TRUE, within=TRUE, hedges.correction=TRUE,conf.level=
0.95,pooled=TRUE)
```

```
##
## Hedges's g (single sample)
##
## g estimate: 0.6067392 (medium)
## Reference mu: 0
## 95 percent confidence interval:
##     lower     upper
## -1.240404  2.453883
```

```
cohen.d(MBB_ses.B_pipeline_results$MeanStrengthaal116despike-MBB_ses_A_pipeline_results$MeanStre
ngthaal116despike, f = NA, paired=TRUE, within=TRUE, hedges.correction=FALSE,conf.level=0.95,poo
led=TRUE)
```

```
##
## Cohen's d (single sample)
##
## d estimate: 0.9605646 (large)
## Reference mu: 0
## 95 percent confidence interval:
##     lower     upper
## -1.662064  3.583194
```

```
cohen.d(MBB_ses.B_pipeline_results$MeanStrengthaal116despike-MBB_ses_A_pipeline_results$MeanStre
ngthaal116despike, f = NA, paired=TRUE, within=TRUE, hedges.correction=TRUE,conf.level=0.95,pool
ed=TRUE)
```

```
##
## Hedges's g (single sample)
##
## g estimate: 0.6985924 (medium)
## Reference mu: 0
## 95 percent confidence interval:
##     lower     upper
## -1.161737  2.558922
```

```
cohen.d(MBB_ses.B_pipeline_results$MeanClusteringCoeffPosaal116despike-MBB_ses_A_pipeline_result
s$MeanClusteringCoeffPosaal116despike, f = NA, paired=TRUE, within=TRUE, hedges.correction=FALSE
,conf.level=0.95,pooled=TRUE)
```

```
##
## Cohen's d (single sample)
##
## d estimate: 1.499557 (large)
## Reference mu: 0
## 95 percent confidence interval:
##     lower     upper
## -1.311197  4.310311
```

```
cohen.d(MBB_ses.B_pipeline_results$MeanClusteringCoeffPosaal116despike-MBB_ses_A_pipeline_result
s$MeanClusteringCoeffPosaal116despike, f = NA, paired=TRUE, within=TRUE, hedges.correction=TRUE,
conf.level=0.95,pooled=TRUE)
```

```
##
## Hedges's g (single sample)
##
## g estimate: 1.090587 (large)
## Reference mu: 0
## 95 percent confidence interval:
##      lower      upper
## -0.8450751  3.0262485
```

### Citations

```
citation(package = "stats")
```

```
##
## The 'stats' package is part of R.  To cite R in publications use:
##
##   R Core Team (2018). R: A language and environment for
##   statistical computing. R Foundation for Statistical Computing,
##   Vienna, Austria. URL https://www.R-project.org/.
##
## A BibTeX entry for LaTeX users is
##
##   @Manual{,
##     title = {R: A Language and Environment for Statistical Computing},
##     author = {{R Core Team}},
##     organization = {R Foundation for Statistical Computing},
##     address = {Vienna, Austria},
##     year = {2018},
##     url = {https://www.R-project.org/},
##   }
##
## We have invested a lot of time and effort in creating R, please
## cite it when using it for data analysis. See also
## 'citation("pkgname")' for citing R packages.
```

```
citation(package = "base")
```

```
##
## To cite R in publications use:
##
##   R Core Team (2018). R: A language and environment for
##   statistical computing. R Foundation for Statistical Computing,
##   Vienna, Austria. URL https://www.R-project.org/.
##
## A BibTeX entry for LaTeX users is
##
##   @Manual{,
##     title = {R: A Language and Environment for Statistical Computing},
##     author = {{R Core Team}},
##     organization = {R Foundation for Statistical Computing},
##     address = {Vienna, Austria},
##     year = {2018},
##     url = {https://www.R-project.org/},
##   }
##
## We have invested a lot of time and effort in creating R, please
## cite it when using it for data analysis. See also
## 'citation("pkgname")' for citing R packages.
```

WSRT:

David F. Bauer (1972). Constructing confidence sets using rank statistics. Journal of the American Statistical Association 67, 687-690. doi: 10.1080/01621459.1972.10481279.

Myles Hollander and Douglas A. Wolfe (1973). Nonparametric Statistical Methods. New York: John Wiley & Sons. Pages 27-33 (one-sample), 68-75 (two-sample). Or second edition (1999).

Patrick Royston (1982). An extension of Shapiro and Wilk's W test for normality to large samples. Applied Statistics, 31, 115-124. doi: 10.2307/2347973.

Shapiro Wilk:

Patrick Royston (1982). Algorithm AS 181: The W test for Normality. Applied Statistics, 31, 176-180. doi: 10.2307/2347986.

Patrick Royston (1995). Remark AS R94: A remark on Algorithm AS 181: The W test for normality. Applied Statistics, 44, 547-551. doi: 10.2307/2986146.

Lesion-Mapper:

lesion_mapper - A tool for mappping white matter hyperintensities. Publication at: Wetter, Hubbard, Motl, Sutton. Brain Behav. 2016 Jan 28;6(3):e00440. https://doi.org/10.1002/brb3.440 (https://doi.org/10.1002/brb3.440)

Brain Connectivity Toolbox:

Complex network measures of brain connectivity: Uses and interpretations. Rubinov M, Sporns O (2010) NeuroImage 52:1059-69.
